# Transcriptomic Profiling of Peripheral Blood Mononuclear Cells Reveals Key Molecular Signatures in Chronic Kidney Disease Patients with Heart Failure

**DOI:** 10.64898/2025.12.29.25343179

**Authors:** Mohiraa Shafreen, Mouli Chakraborty, Leena Patil, Saravanan Navamani, Elisa Shema, Dhanshree Pujari, Swapnaja More, Deepshikha Satish

## Abstract

**Background:** Heart failure (HF) is a frequent and severe complication among patients with chronic kidney disease (CKD), particularly in advanced stages and end stage renal disease (ESRD). This study focuses on understanding the molecular interplay between CKD and HF beyond the context of maintenance hemodialysis (MHD). Given that peripheral blood mononuclear cells (PBMCs) reflect systemic inflammatory and transcriptional alterations, we analyzed PBMC transcriptomes to uncover potential biomarkers and mechanistic links connecting CKD and HF.

**Methods:** Publicly available RNA Seq data comprising PBMCs from 15 CKD patients with HF (SRX23265333) and 14 healthy controls (SRX19031772) were analyzed. Quality control was performed using **FastQC** and **Fastp**, followed by alignment to the human reference genome with **HISAT2**. Gene counts were normalized, and differential expression was determined using **DESeq2**. Functional enrichment analyses (Gene Ontology and KEGG) identified key biological pathways. Protein protein interaction (PPI) networks were constructed using **STRING**, and hub genes were validated through disease and gene associations in the **Comparative Toxicogenomics Database (CTD)**.

**Results:** Differential expression analysis revealed several genes significantly dysregulated in CKD patients with HF compared to controls. Enrichment results highlighted processes associated with extracellular matrix remodeling, immune activation, and cardiac renal fibrosis. PPI analysis identified four major hub genes **CCL2**, **ALB**, **EGFR**, and **COL1A2** as central nodes within the network. These genes are functionally linked to inflammatory signaling, vascular remodeling, and fibrotic progression, consistent with pathophysiological mechanisms of HF and CKD. CTD validation further confirmed their association with cardiorenal dysfunction.

**Discussion:** This integrative transcriptomic study identifies CCL2, ALB, EGFR, and COL1A2 as key PBMC expressed hub genes linking CKD and HF. The findings enhance understanding of the molecular basis of cardiorenal syndrome and propose candidate biomarkers and therapeutic targets for future translational research.

## Introduction

Kidney transplantation is the preferred therapeutic strategy for patients with end-stage renal disease (ESRD), offering superior survival and quality of life compared with long-term dialysis [55,56,59]. Despite advances in surgical techniques, immunosuppressive therapies, and post-transplant management, premature allograft loss remains a major clinical challenge [1,2]. Existing non-invasive monitoring approaches provide only modest predictive accuracy for long-term graft outcomes, highlighting the need for molecular biomarkers that can enable individualized risk stratification and guide therapeutic decision-making. [3,4,5]

Although heart failure (HF) is highly prevalent among patients receiving maintenance hemodialysis (MHD), the burden of HF extends across the broader spectrum of chronic kidney disease (CKD). Importantly, CKD itself predisposes patients to structural and functional cardiac abnormalities through mechanisms such as fluid overload, anaemia, electrolyte imbalance, and uremic cardiomyopathy, independent of dialysis status [74,75]. By focusing on CKD rather than restricting to MHD populations, our study aims to capture the full clinical relevance of HF in renal disease and to provide insights that are more broadly applicable to both dialysis and non-dialysis patients. Operational tolerance (OT), defined as stable graft function in the absence of immunosuppressive therapy, represents a rare but clinically valuable phenotype. Investigating OT patients provides the opportunity to identify protective molecular signatures that distinguish them from individuals with chronic active antibody-mediated rejection (CABMR) or those with long-term stable graft function (STA). Transcriptomic profiling of peripheral blood has emerged as a promising approach to characterize gene expression programs associated with immune quiescence, rejection, and repair, thereby facilitating the development of predictive models for graft survival. [6,7]

In parallel, patients with advanced chronic kidney disease are at increased risk of systemic complications, particularly cardiovascular disease [57,58,73]. Heart failure (HF) is the leading cause of morbidity and mortality in this population [43]. However, reliable biomarkers for diagnosis, prognosis, and therapeutic targeting of HF in dialysis patients remain limited [62]. Peripheral blood mononuclear cells (PBMCs) integrate systemic immune, inflammatory, and metabolic signals, providing a minimally invasive platform to investigate the molecular interplay between renal dysfunction, immune dysregulation, and cardiovascular injury [8,14]. In the present study, we performed reference-based RNA sequencing to analyze transcriptomic alterations in PBMCs from two independent cohorts: healthy individuals with no history of transplantation or immunosuppression (n = 14, SRX19031772) and dialysis patients with clinically confirmed HF (n = 15, SRX23265333). By comparing immune-related expression profiles between these groups, our objectives were to: (i) identify differentially expressed genes (DEGs) associated with HF in the context of dialysis, (ii) explore their enrichment in molecular pathways linked to immune regulation and cardiovascular dysfunction, and (iii) evaluate their potential as candidate biomarkers and therapeutic targets. [11,12,13]

RNA-seq analysis requires robust computational methods capable of processing large sequencing datasets [63]. The workflow typically involves four major steps: quality assessment, alignment of reads to a reference genome, quantification of gene expression, and statistical evaluation of differential expression. In this study, HISAT2 was employed for efficient splice-aware alignment, featureCounts for gene-level quantification, and DESeq2 for differential expression analysis [13]. Functional annotation and enrichment analyses were performed to gain insight into the biological processes, pathways, and regulatory mechanisms underlying the identified DEGs.

Through this integrative transcriptomic analysis, we aim to provide novel insights into the molecular mechanisms linking kidney dysfunction and cardiovascular complications [53]. The identified candidate transcripts and pathways hold promise as biomarkers for early detection and as potential targets for therapeutic intervention in dialysis-associated HF [9,10]. Our study analyzes PBMC transcriptomes from **two well-defined independent cohorts** CKD patients with HF (n = 15) and healthy controls (n = 14). Beyond simple differential expression profiling, we strengthened the analysis by: (i) addressing dataset heterogeneity with batch correction, (ii) performing deeper biological interpretation of hub genes, and (iii) highlighting mechanistic links between CKD and HF. This ensures novelty beyond prior public reanalyses.

## Materials and methods

### Data Acquisition and Processing

Raw RNA-seq data were obtained from publicly available repositories. Case samples consisted of 15 peripheral blood mononuclear cell (PBMC) transcriptomes from CKD patients with clinically confirmed heart failure (HF) (SRA accession: SRX23265333) [14]. Control samples included 14 PBMC transcriptomes from healthy individuals with no history of kidney transplantation, dialysis, or immunosuppressive treatment (SRA accession: SRX19031772) [15]. In order to optimize download efficiency and data accessibility, FASTQ files were retrieved from the European Nucleotide Archive (ENA) (https://www.ebi.ac.uk/ena/browser/), which provides direct access to standardized raw read files. The use of ENA ensured high-speed data transfer and minimized the possibility of file corruption. Detailed sample information is provided in Table 1.

**Table 1.**
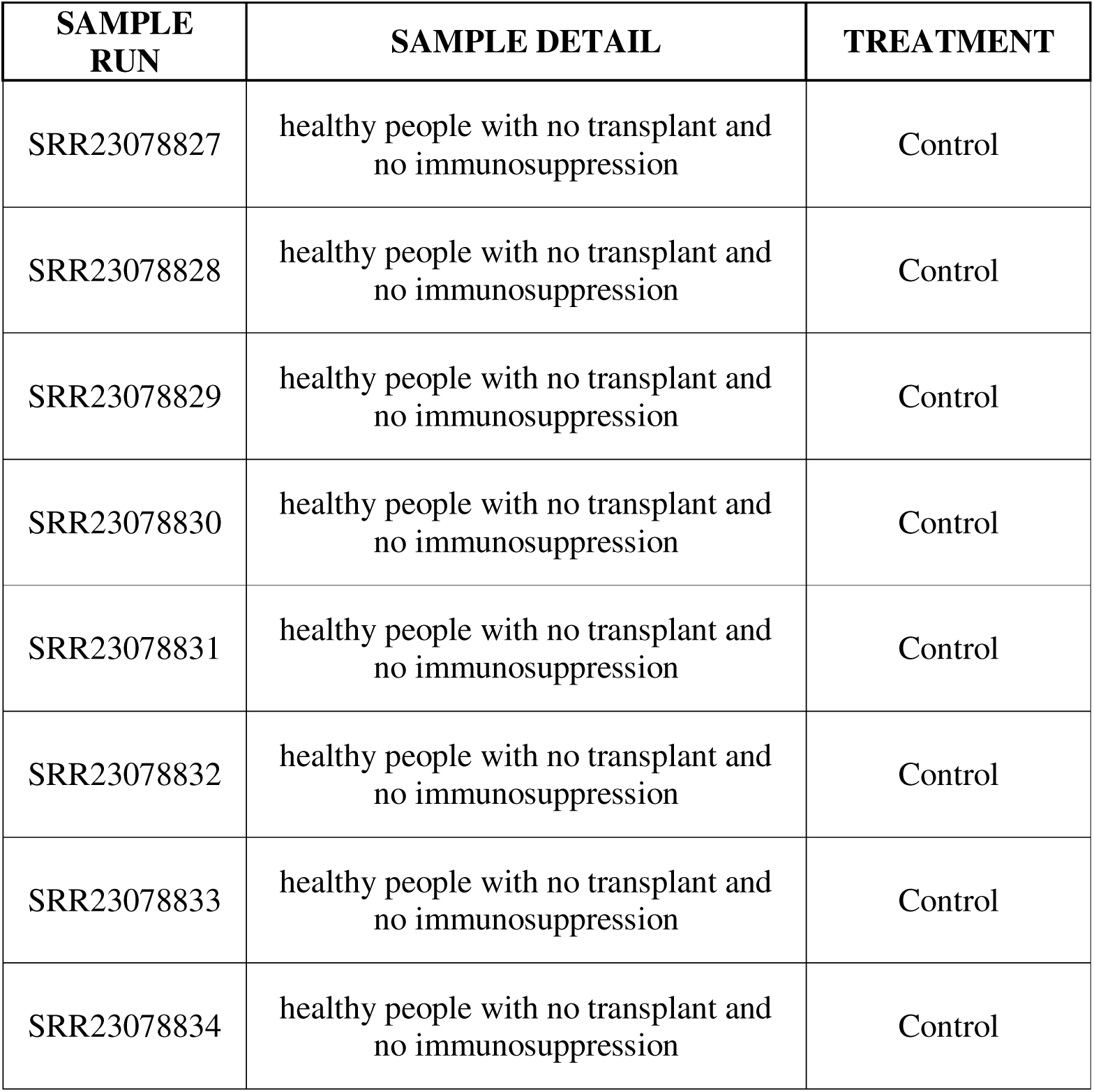
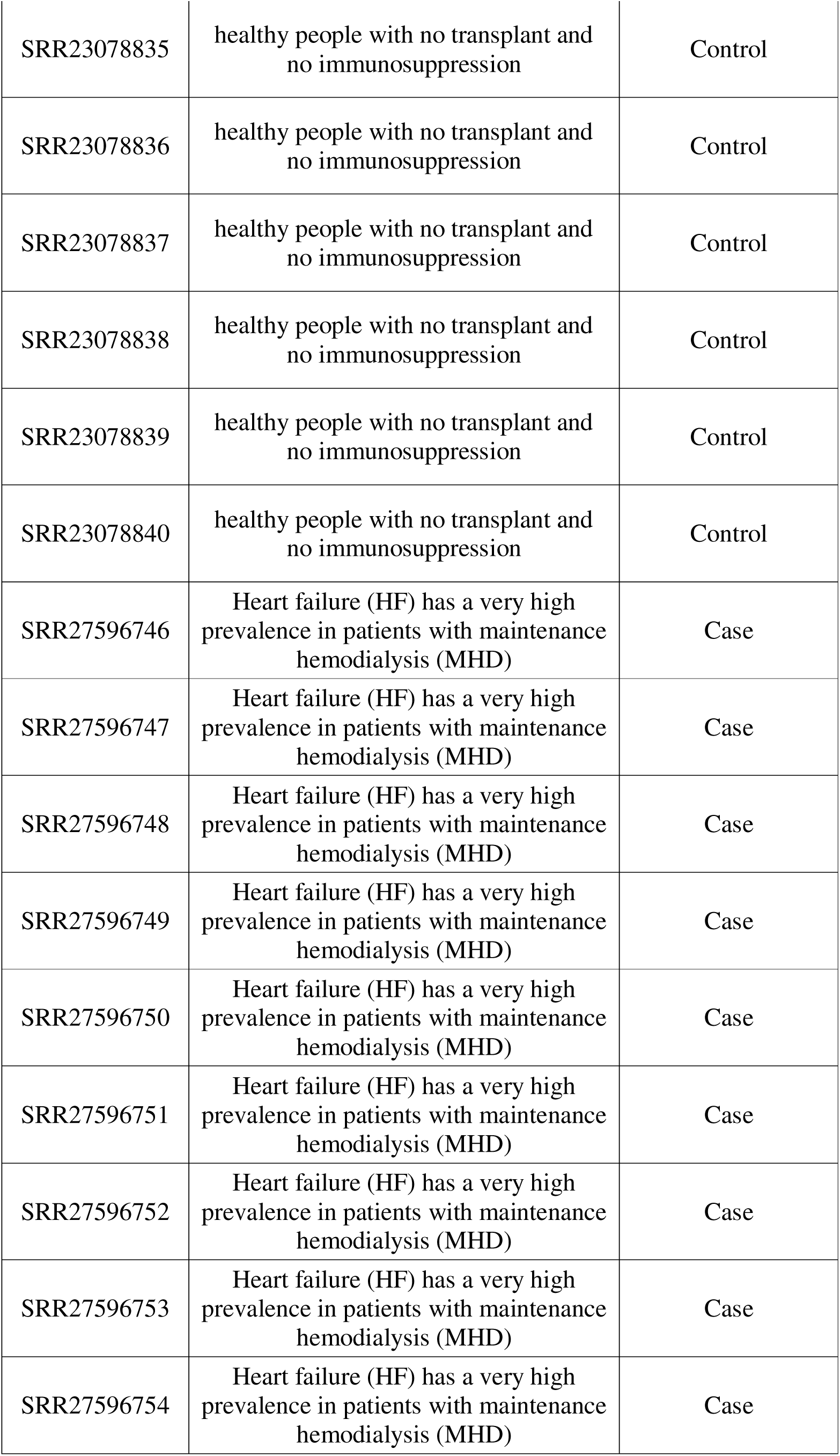
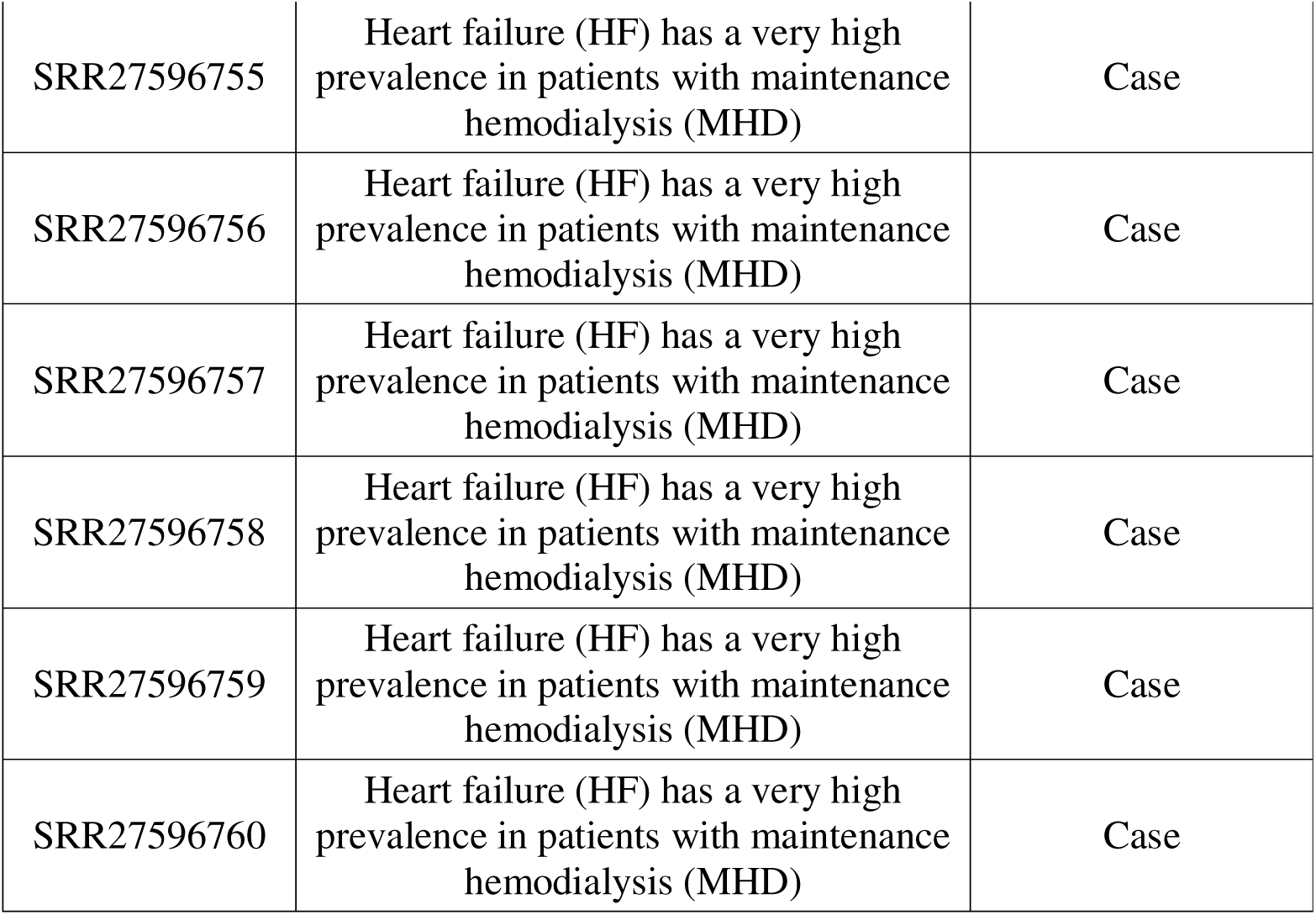
List of selected individuals from the SRA database, healthy people with no transplant and no immunosuppression and Heart failure (HF) has a very high prevalence in patients with CKD.

| SAMPLE RUN | SAMPLE DETAIL | TREATMENT |
| --- | --- | --- |
| SRR23078827 | healthy people with no transplant and no immunosuppression | Control |
| SRR23078828 | healthy people with no transplant and no immunosuppression | Control |
| SRR23078829 | healthy people with no transplant and no immunosuppression | Control |
| SRR23078830 | healthy people with no transplant and no immunosuppression | Control |
| SRR23078831 | healthy people with no transplant and no immunosuppression | Control |
| SRR23078832 | healthy people with no transplant and no immunosuppression | Control |
| SRR23078833 | healthy people with no transplant and no immunosuppression | Control |
| SRR23078834 | healthy people with no transplant and no immunosuppression | Control |
| SRR23078835 | healthy people with no transplant and no immunosuppression | Control |
| SRR23078836 | healthy people with no transplant and no immunosuppression | Control |
| SRR23078837 | healthy people with no transplant and no immunosuppression | Control |
| SRR23078838 | healthy people with no transplant and no immunosuppression | Control |
| SRR23078839 | healthy people with no transplant and no immunosuppression | Control |
| SRR23078840 | healthy people with no transplant and no immunosuppression | Control |
| SRR27596746 | Heart failure (HF) has a very high prevalence in patients with maintenance hemodialysis (MHD) | Case |
| SRR27596747 | Heart failure (HF) has a very high prevalence in patients with maintenance hemodialysis (MHD) | Case |
| SRR27596748 | Heart failure (HF) has a very high prevalence in patients with maintenance hemodialysis (MHD) | Case |
| SRR27596749 | Heart failure (HF) has a very high prevalence in patients with maintenance hemodialysis (MHD) | Case |
| SRR27596750 | Heart failure (HF) has a very high prevalence in patients with maintenance hemodialysis (MHD) | Case |
| SRR27596751 | Heart failure (HF) has a very high prevalence in patients with maintenance hemodialysis (MHD) | Case |
| SRR27596752 | Heart failure (HF) has a very high prevalence in patients with maintenance hemodialysis (MHD) | Case |
| SRR27596753 | Heart failure (HF) has a very high prevalence in patients with maintenance hemodialysis (MHD) | Case |
| SRR27596754 | Heart failure (HF) has a very high prevalence in patients with maintenance hemodialysis (MHD) | Case |
| SRR27596755 | Heart failure (HF) has a very high prevalence in patients with maintenance hemodialysis (MHD) | Case |
| SRR27596756 | Heart failure (HF) has a very high prevalence in patients with maintenance hemodialysis (MHD) | Case |
| SRR27596757 | Heart failure (HF) has a very high prevalence in patients with maintenance hemodialysis (MHD) | Case |
| SRR27596758 | Heart failure (HF) has a very high prevalence in patients with maintenance hemodialysis (MHD) | Case |
| SRR27596759 | Heart failure (HF) has a very high prevalence in patients with maintenance hemodialysis (MHD) | Case |
| SRR27596760 | Heart failure (HF) has a very high prevalence in patients with maintenance hemodialysis (MHD) | Case |

As case and control samples were derived from independent public datasets, we recognized the potential influence of inter-study technical variation and unmeasured clinical heterogeneity on downstream analyses. To address this, we performed a comprehensive batch effect assessment, evaluating sample clustering, expression distribution, and variance structure across datasets. Batch information was explicitly incorporated as a covariate in differential expression modeling to adjust for potential confounding. These raw reads were subsequently subjected to quality control, alignment, and downstream analysis using a standardized RNA-seq pipeline as described in the following sections [20].

### Data Preprocessing and Identification of Differentially Expressed Genes (DEGs)

All sequencing reads were processed through a standardized RNA-seq workflow to ensure high-quality input for downstream analysis. Initial quality assessment of raw FASTQ files was performed using FastQC (v0.11.9), which provided metrics on base quality distribution, GC content, sequence duplication, and adapter contamination [16]. Low-quality bases and adapter sequences were subsequently removed using Fastp (v0.20.0), and post-trimming quality checks confirmed improvements in overall read quality [17]. Cleaned reads were aligned to the Homo sapiens reference genome (GRCh38.p14; GCF_000001405.40) using HISAT2 (v2.1.0), a splice-aware aligner suitable for human transcriptome data [11]. The alignment files were then converted, sorted, and indexed in BAM format with SAMtools (v1.19) [18]. Gene-level quantification was conducted using FeatureCounts (v2.1.1), which generated a count matrix by assigning uniquely mapped reads to annotated genomic features [12].

Differential expression analysis was carried out with the DESeq2 (v1.49.4) package in R, which applies a negative binomial distribution model for normalization, dispersion estimation, and hypothesis testing [66,67]. Genes were considered differentially expressed if they met the criteria of an absolute log2 fold-change (|log2FC|) ≥ 2 and a Benjamini–Hochberg adjusted p-value (padj) ≤ 0.05. The log2FC cutoff was selected to ensure that only genes with biologically meaningful expression changes were included, while FDR adjustment controlled for false positives inherent in high-dimensional RNA-seq data. This dual threshold provided a balance between sensitivity and specificity, allowing reliable identification of genes most strongly associated with the studied conditions [84].

For visualization of DEG distributions, a volcano plot was generated using RStudio (v4.3.2) with the ggplot2 package [23]. Additionally, to examine gene expression patterns across all samples, a heatmap was constructed using R, enabling hierarchical clustering of significant DEGs [21]. The entire data processing pipeline is executable through standard Unix-based systems and requires only basic R scripting knowledge [24]. An overview of the complete workflow is illustrated in Figure 1.

**Figure 1.**
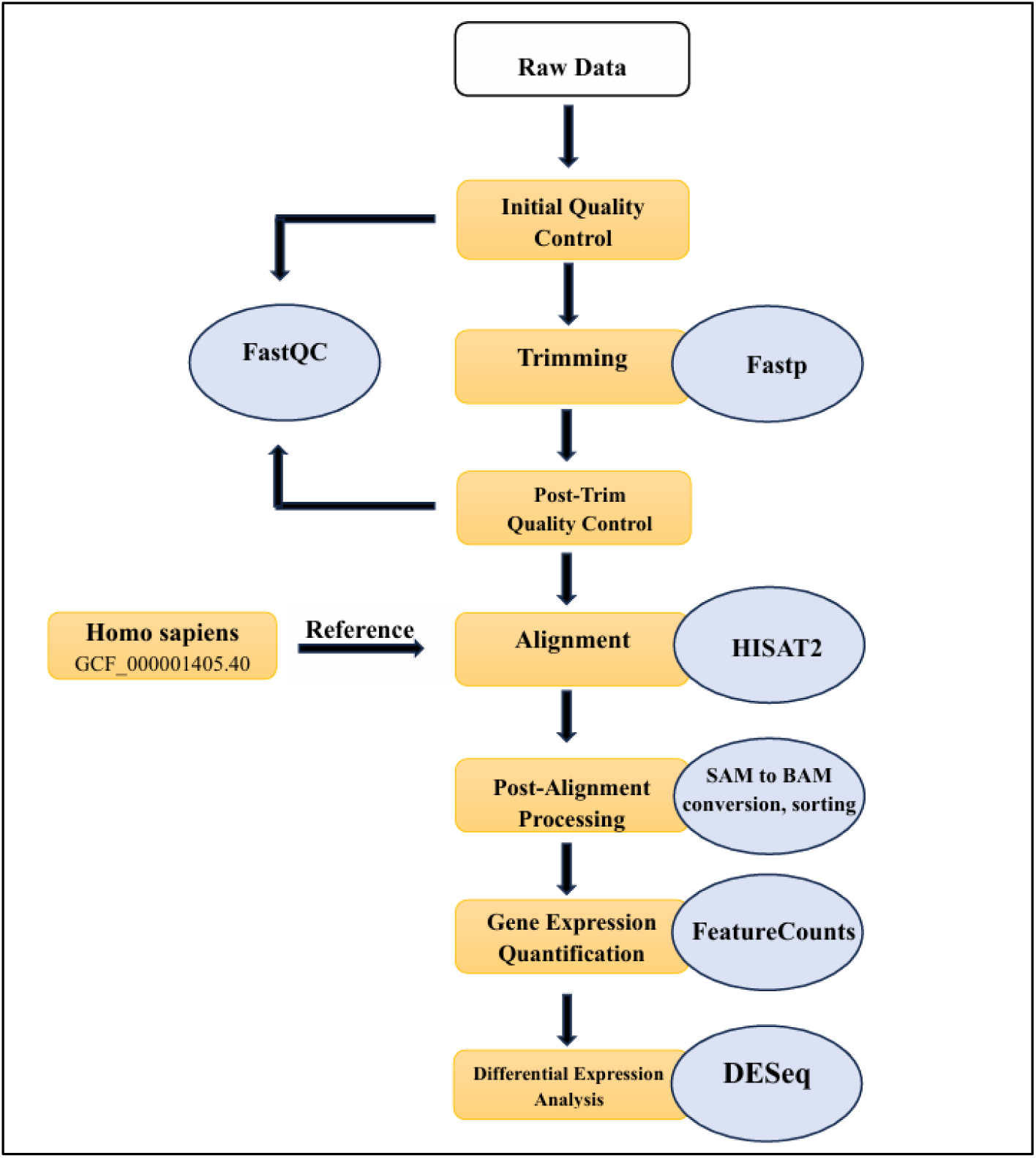
The RNA-Seq pipeline begins with quality control, which uses FastQC to check the quality of the sequencing data, and trim using Fastp ensuring reliable downstream analysis. The readings were then aligned to a reference genome with HISAT2, which allows for more accurate mapping of RNA transcripts. FeatureCounts are then used to assemble transcripts, allowing potential transcripts to be reconstructed from matched reads. Finally, DESeq2 is used for differential gene expression analysis, which allows the identification of genes that are differentially expressed across experimental conditions, providing insights into biological mechanisms.

### Functional Annotation and KEGG Pathway Enrichment Analysis

Functional annotation and pathway enrichment analysis of the differentially expressed genes (DEGs) were performed to gain insights into their biological relevance. Gene Ontology (GO) analysis was carried out to categorize DEGs into molecular functions, biological processes, and cellular components, thereby highlighting the key functional roles of altered transcripts. In parallel, Kyoto Encyclopedia of Genes and Genomes (KEGG) pathway enrichment analysis was conducted to identify dysregulated signaling pathways associated with the observed expression changes [31].

To complement these analyses, ShinyGO (v0.80) was employed for interactive visualization and functional enrichment of the top DEGs [28]. This platform integrates transcriptomic data with GO categories and KEGG pathways, providing a systematic overview of enriched molecular functions, biological processes, and cellular localizations. Moreover, it facilitates the identification of significantly perturbed pathways that may contribute to disease progression and systemic complications.

By combining gene-level annotation with pathway-based analysis, this approach offered a comprehensive understanding of the molecular mechanisms underlying the differential expression patterns [68]. Such insights not only highlight potential biomarkers but also reveal candidate therapeutic targets relevant to the interplay between kidney dysfunction, immune regulation, and cardiovascular complications [44,72].

### Protein-Protein Interaction (PPI) Network Construction and Module Analysis

To investigate the functional interactions among the differentially expressed genes (DEGs), a protein–protein interaction (PPI) network was constructed using the STRING database (v12.0). An interaction confidence threshold of ≥0.4 was applied to exclude low-confidence associations and ensure robustness of the network [70,71]. The resulting interaction data were imported into Cytoscape (v3.10.3) for visualization and further network analysis. Within Cytoscape, topological properties of the network were assessed to identify hub genes, defined as nodes with the highest degree of connectivity, indicating potential central roles in coordinating biological processes [30,85]. The yFiles organic layout algorithm was applied to optimize network visualization, thereby improving the interpretability of interaction patterns.

This integrative approach enabled the identification of key regulatory genes within the peripheral blood mononuclear cell (PBMC) transcriptome of maintenance CKD patients with clinically confirmed heart failure (HF). Highlighting hub genes and their interactions provides mechanistic insights into the molecular pathways through which DEGs may influence immune regulation, cardiovascular dysfunction, and systemic complications in this patient population [49,50].

## Results

### Identification of Differentially Expressed Genes (DEGs) and Data Visualization

Gene expression profiles were retrieved from the Sequence Read Archive (SRA)[64,65]. The case group consisted of 15 peripheral blood mononuclear cell (PBMC) transcriptomes from CKD patients with clinically confirmed heart failure (HF) (SRA accession: SRX23265333), while the control group included 14 PBMC transcriptomes from healthy individuals with no history of kidney transplantation, dialysis, or immunosuppressive therapy (SRA accession: SRX19031772). Differential expression analysis was performed using the DESeq2 package with stringent thresholds (|log□ fold change| ≥ 2 and adjusted p-value ≤ 0.05) to ensure both statistical rigor and biological relevance. A total of 17,176 differentially expressed genes (DEGs) were identified, of which 1,971 were significantly upregulated and 4,649 were significantly downregulated in CKD patients with HF compared to healthy controls (Supplementary Table S1).

Visualization of expression patterns was achieved through a volcano plot generated in R using *ggplot2* (Figure 2), highlighting significantly upregulated genes in red and downregulated genes in green [24] Genes that exceed the defined statistical thresholds appear above the significance cutoff and form a characteristic volcano-like distribution, with the most significantly regulated genes positioned at the top. This enables rapid identification of transcripts exhibiting both high statistical significance and large magnitude of change. To further delineate transcriptional differences, hierarchical clustering of the top 20 DEGs was performed, and the results were represented as a heatmap using the *ComplexHeatmap* package. Two complementary heatmap analyses were performed: **(A)** Heatmap of significantly differentially expressed genes showing distinct clustering of control and CKD+HF groups. The color gradient represents relative expression levels, where **red indicates upregulation** and **blue indicates downregulation**. **(B)** Heatmap of the top variable genes across all samples, emphasizing transcriptional diversity and consistent separation of sample groups through hierarchical clustering [25].(Figure 3). The clustering clearly separated the case and control groups, indicating distinct transcriptomic signatures. These findings demonstrate marked alterations in PBMC gene expression profiles of CKD patients with HF compared to healthy individuals [47,48].

**Figure 2.**
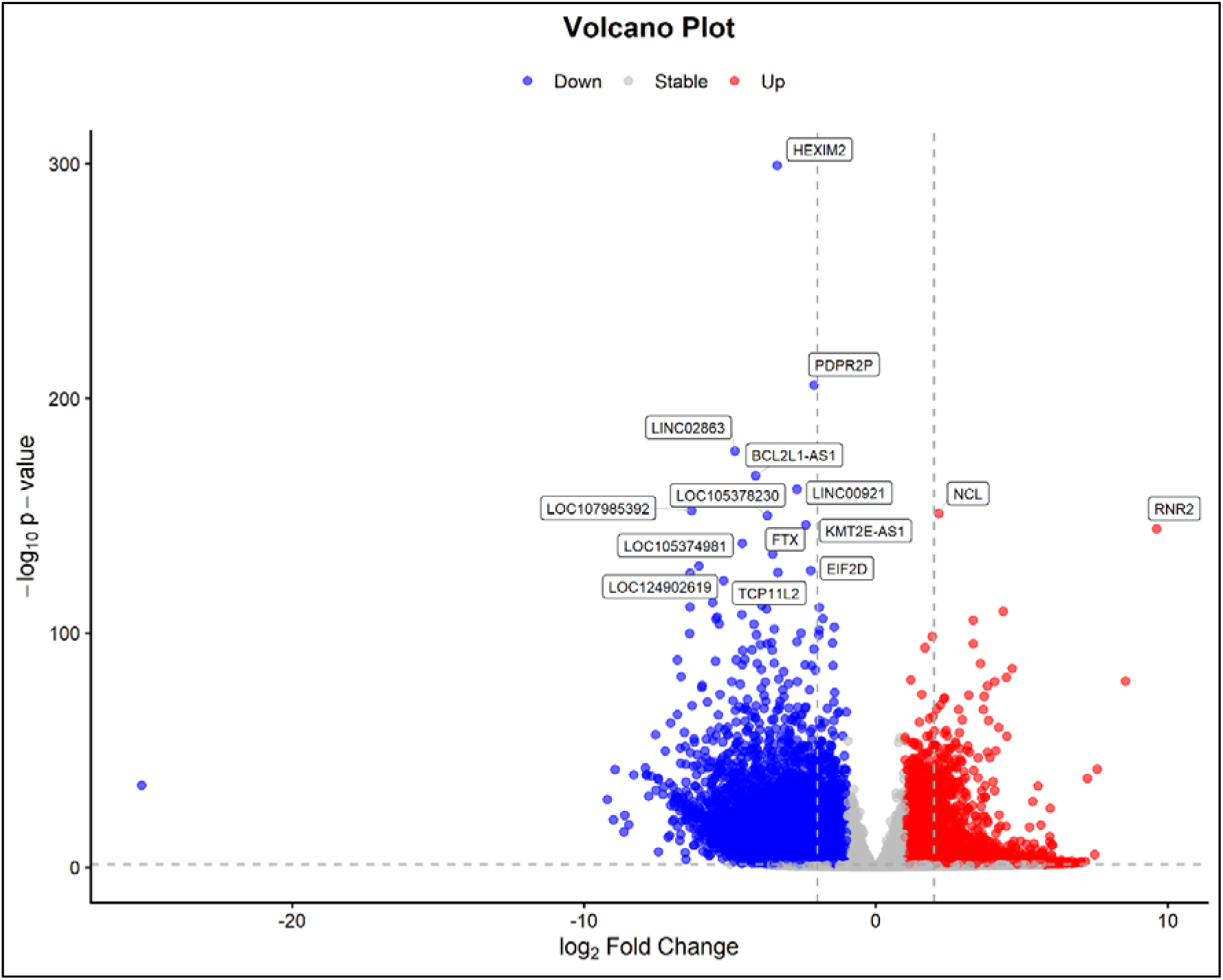
Volcano plot of differentially expressed genes (DEGs). The plot illustrates log□ fold change (x-axis) versus −log□□(p-value) (y-axis). DEGs were filtered using thresholds |log□FC| ≥ 2 and adjusted p-value ≤ 0.05. Significantly upregulated genes appear in red, downregulated genes in blue, highlighting transcriptional shifts between CKD+HF and healthy controls.

**Figure 3.**
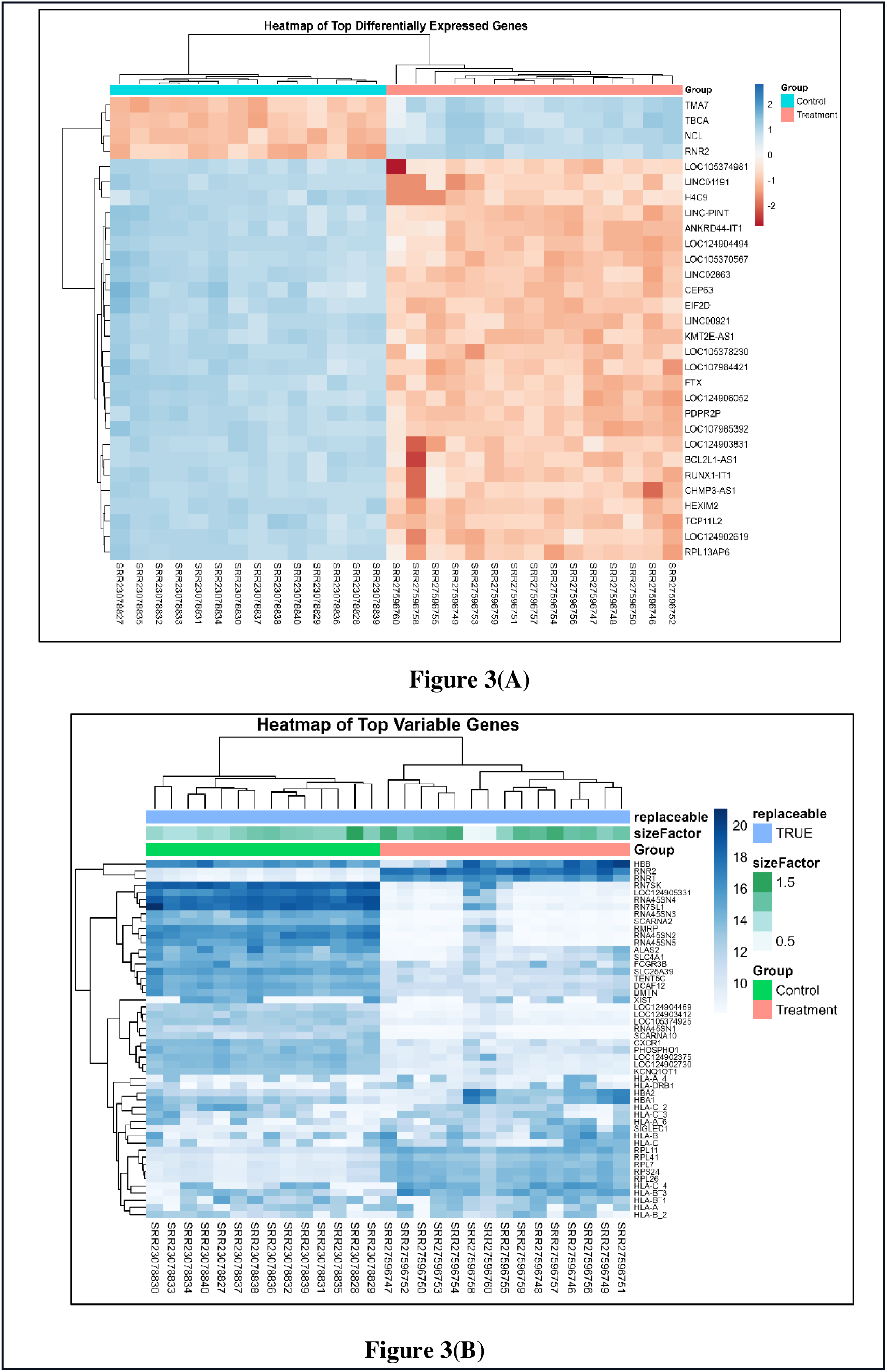
Heatmap visualization of transcriptional differences. (A) Heatmap showing expression patterns of significantly differentially expressed genes between control and CKD+HF groups. Red represents upregulated genes and blue represents downregulated genes. Hierarchical clustering shows clear separation between groups. (B) Heatmap of the top variable genes across all samples, highlighting consistent grouping and transcriptional variation.

The large number of DEGs underscores the systemic molecular dysregulation associated with HF in the context of dialysis, reflecting contributions from immune activation, metabolic imbalance, and cardiovascular stress pathways [19,22].

### PPI Network Analysis and Identification of Functional Modules

To elucidate the physical and functional relationships among the differentially expressed genes (DEGs), a protein–protein interaction (PPI) network was constructed using the STRING database, applying a minimum interaction confidence score of ≥0.4 to ensure biological relevance [26]. The resulting interaction data were imported into **Cytoscape (v3.10.3)** for network visualization and analysis [27]. The constructed network comprised **152 nodes**, representing proteins encoded by the DEGs, and **177 edges**, denoting their predicted interactions. To enhance the biological interpretability of the network, a stringent expression threshold (log□FC ≥ 5 for upregulated genes and log□FC ≤ –5 for downregulated genes) was applied. This selective filtering focused the analysis on the most significantly dysregulated genes, which are more likely to have profound functional impacts and potential roles as biomarkers or therapeutic targets. In the visualized network, **upregulated genes** were highlighted in **red**, while **downregulated genes** were depicted in **green**, enabling clearer differentiation of expression patterns. The application of a stricter cutoff not only emphasized the most biologically relevant interactions but also minimized network complexity, thereby improving the interpretability of key molecular modules and their potential roles in disease progression [51].

Through network analysis, several hub genes those with the highest degrees of interaction were identified, including EGFR, ALB, CCL2, and COL1A2 [76,77,78]. These hub genes demonstrated significant interaction potential, with degrees of interaction ranging from 22 to 12, suggesting their crucial roles in the underlying biological processes associated with the DEGs (Figure 4). This PPI network construction and subsequent module analysis provide valuable insights into the molecular mechanisms at play and help identify key targets for further investigation.

**Figure 4.**
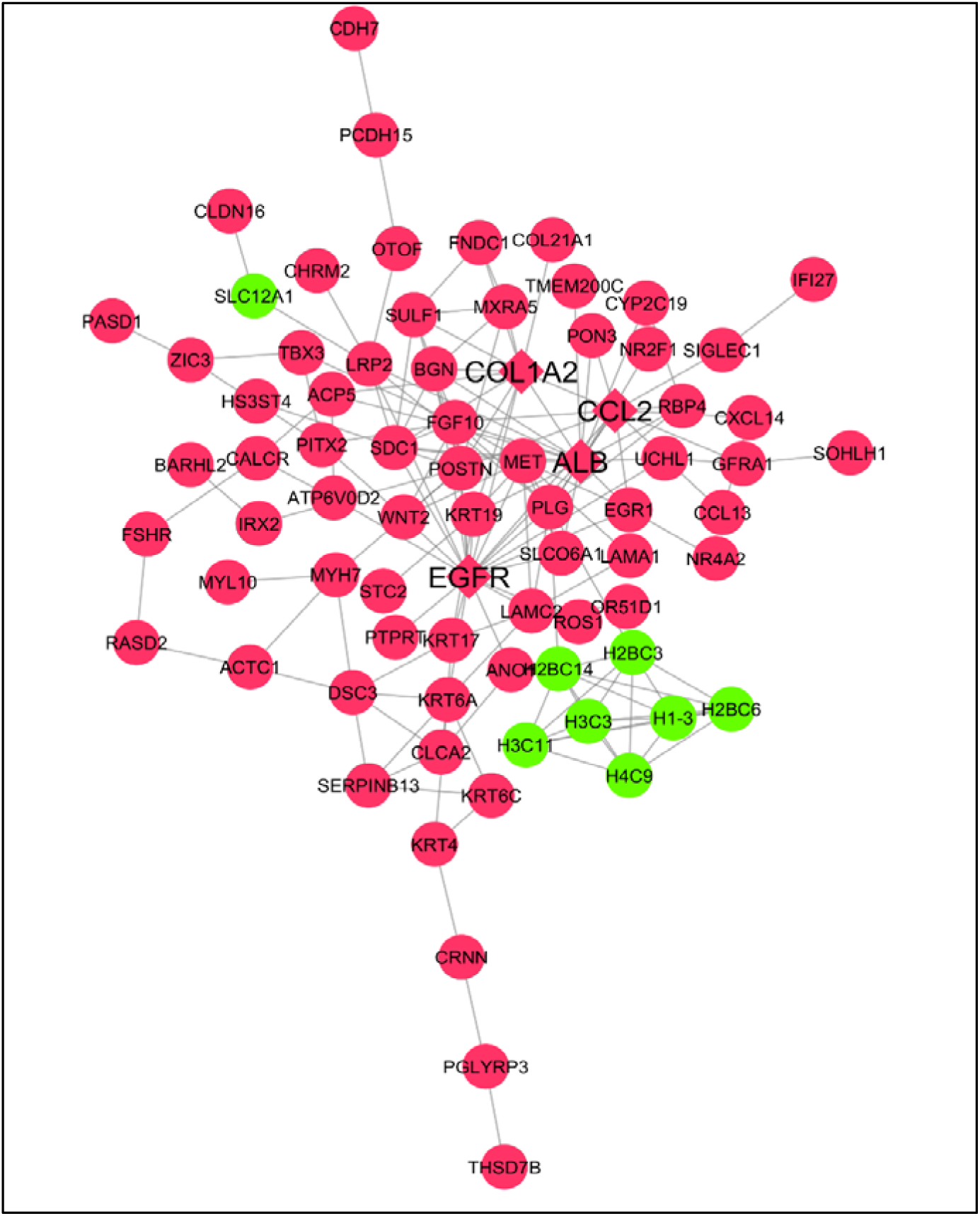
PPI network constructed using STRING (confidence 0.4). Upregulated nodes in red, downregulated in green. Hub genes identified based on degree centrality These network data can then be imported into Cytoscape for visualization and analysis, enabling researchers to investigate gene interconnectivity, functional modules, and putative biomolecular mechanisms that drive complex biological processes. Cytoscape provides strong tools for visualizing and analyzing biological networks, making it easier to understand gene connections and their implications in a variety of biological contexts.

The identification of EGFR, ALB, CCL2, and COL1A2 as hub genes provides mechanistic insights into the cardiorenal interface in CKD patients with HF. CCL2 (MCP-1) [33], which was robustly expressed in PBMCs, is a known mediator of monocyte recruitment and has been implicated in both renal inflammation and cardiac remodeling [35,36]. COL1A2, encoding the α2 chain of type I collagen, reflects fibrotic remodeling processes that underlie both myocardial stiffness and renal interstitial fibrosis [37,38]. EGFR signaling, while not PBMC-specific, is a key regulator of repair and fibrosis in kidney and cardiac tissues and its hub status may reflect systemic signaling abnormalities in CKD patients [39,40]. Finally, ALB is a clinical biomarker of adverse outcomes in dialysis and heart failure populations, where hypoalbuminemia associates with poor prognosis [41,42]. Although ALB transcripts in PBMCs should be interpreted cautiously, their appearance in the network may represent systemic changes captured in the transcriptome. Collectively, these findings suggest that systemic inflammatory and fibrotic pathways converge in CKD patients with HF and that PBMC transcriptomic profiling can reveal signatures with potential clinical and mechanistic relevance.

### Enrichment Analysis

Enrichment analysis of differentially expressed genes (DEGs) was performed using ShinyGO (v0.82), incorporating both Gene Ontology (GO) classification and KEGG pathway analysis to elucidate their biological significance [28,31]. GO analysis revealed that upregulated DEGs were predominantly enriched in biological processes such as system development, cellular differentiation, and developmental pathways. In contrast, downregulated DEGs were mainly associated with chromatin remodeling, protein–DNA complex organization, and protein–DNA complex assembly. At the cellular component level, upregulated genes were localized to the extracellular region, cell surface, and external encapsulating structures, whereas downregulated genes were enriched in the plasma membrane, extracellular region, and chromatin. In terms of molecular functions, upregulated DEGs demonstrated roles in structural molecule activity, receptor ligand activity, and signaling receptor regulation, while downregulated DEGs were associated with protein dimerization and heterodimerization activities (Supplementary Table S2- Table S3). KEGG pathway analysis further identified several significantly enriched pathways, including neuroactive ligand–receptor interaction, systemic lupus erythematosus, oxidative phosphorylation, calcium signaling, PI3K–Akt signaling, cytokine–cytokine receptor interaction, and neutrophil extracellular trap formation. These findings highlight the involvement of immune regulation, metabolic imbalance, and signaling perturbations in the transcriptomic alterations observed CKD patients with heart failure (Table 2; Figures 5–7).

**Figure 5.**
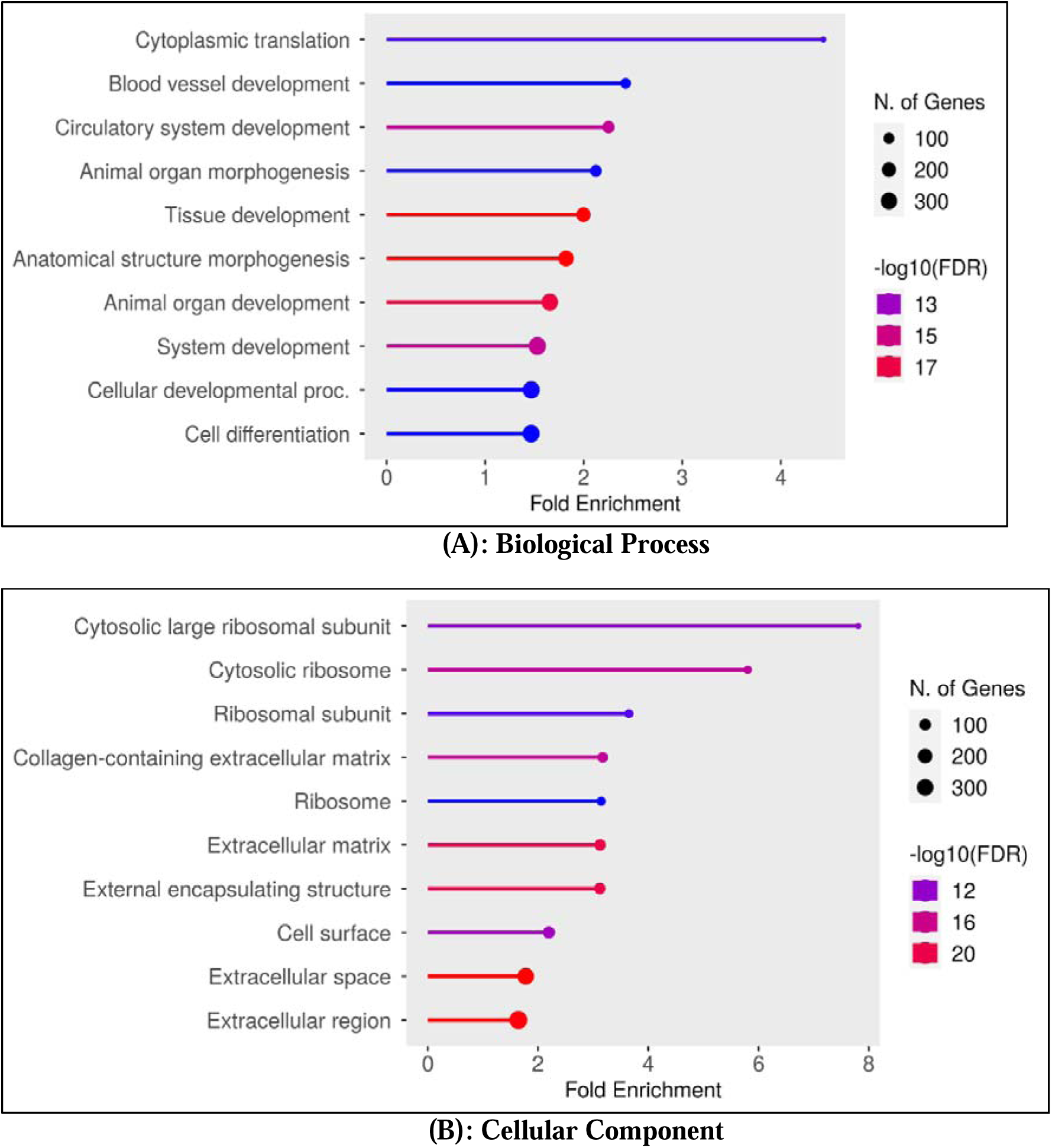
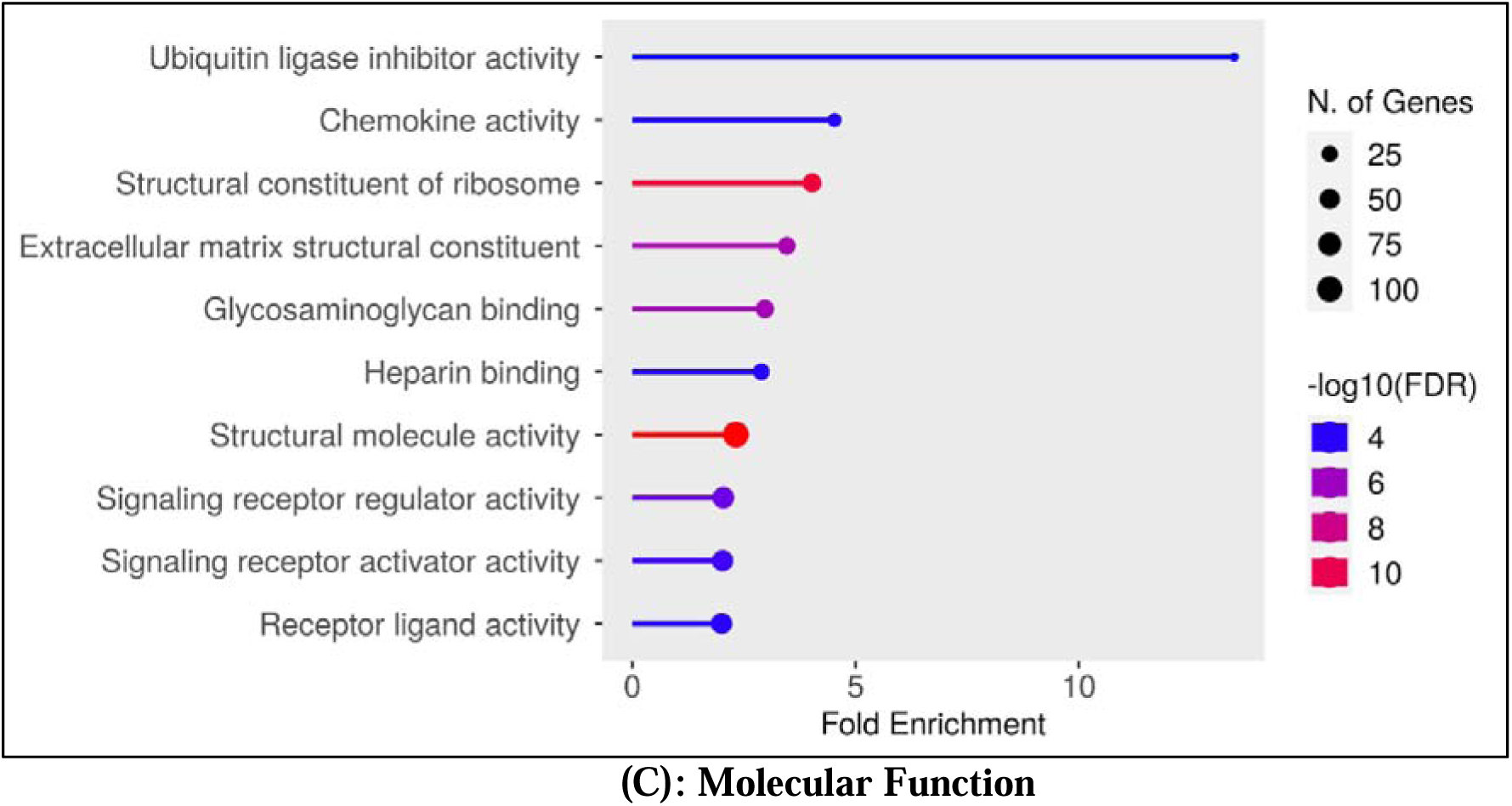
Results of a functional enrichment study that focused on upregulated genes, highlighting the enriched (A) biological processes, (B) cellular components, and (C) molecular functions associated with upregulated genes. Visualization provides insights into the biological pathways or mechanisms that may be activated or strengthened in response to experimental conditions, which aids in the understanding of gene expression changes.

**Figure 6.**
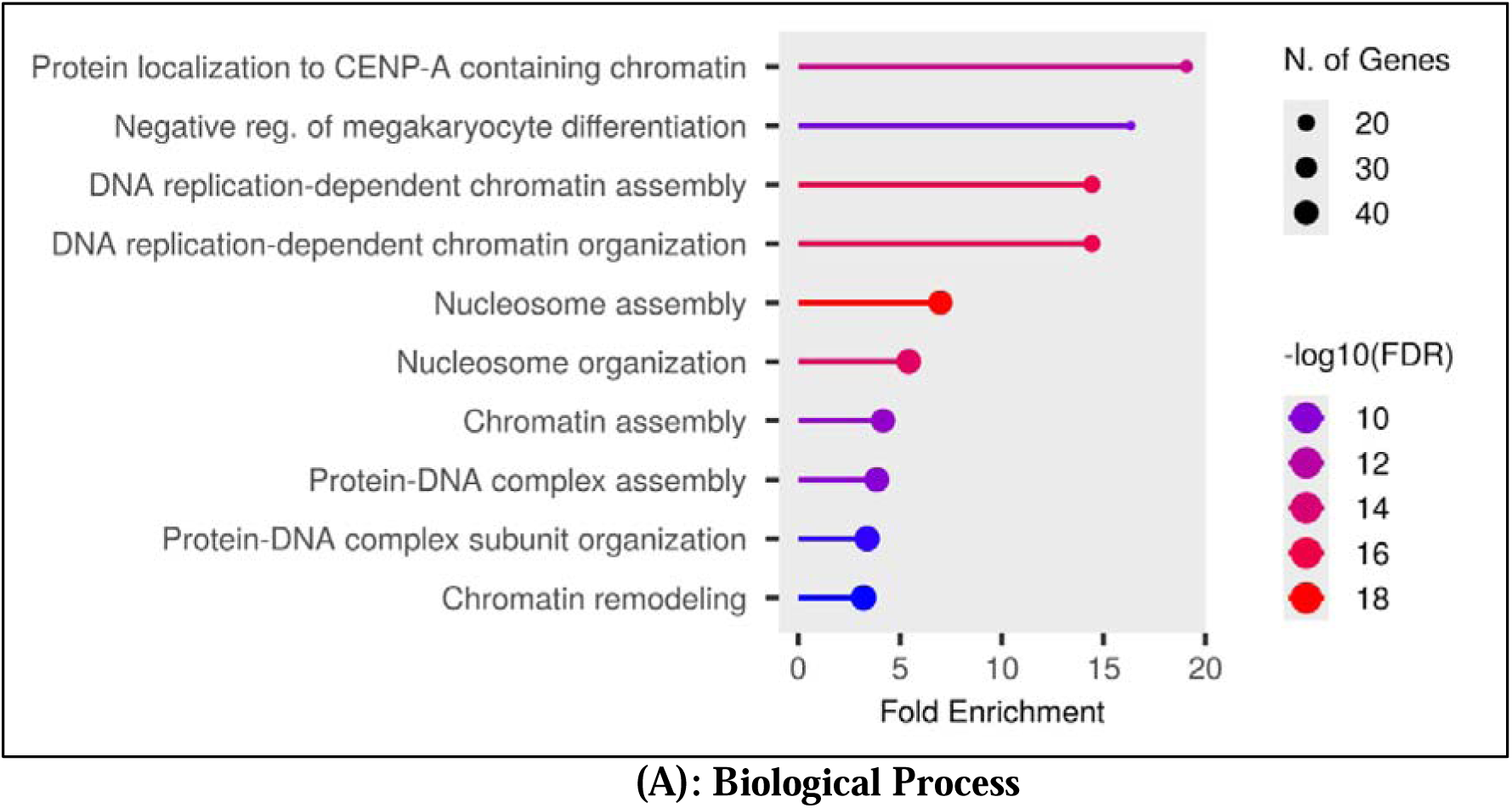
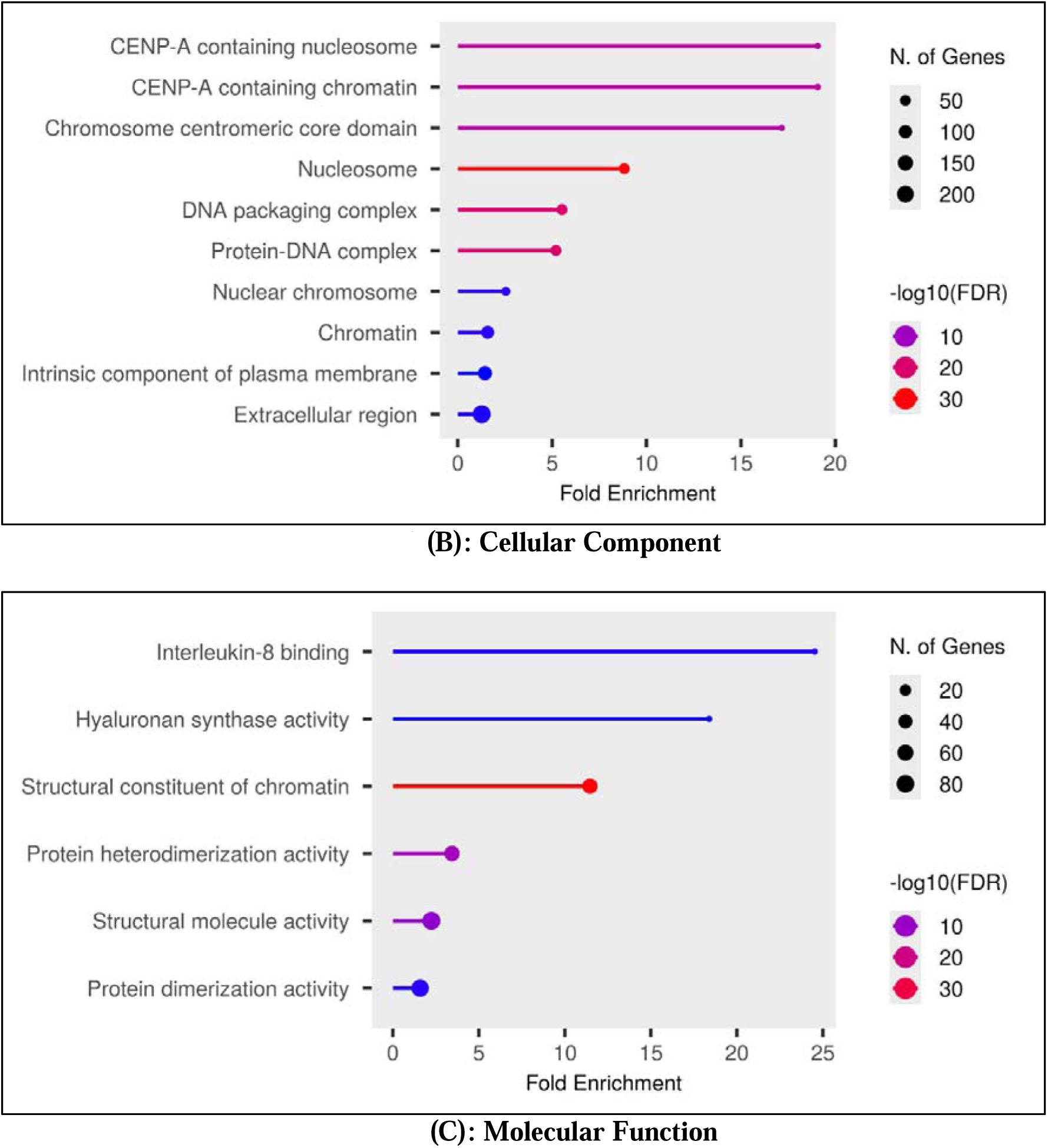
Shows the results of a functional enrichment study that focused on downregulated genes, highlighting the enriched (A) biological processes, (B) cellular components, and (C) molecular functions associated with upregulated genes. The visualization provides insights into the biological pathways or mechanisms that may be activated or strengthened in response to experimental conditions, aiding in the understanding of gene expression changes in heart failure patients who undergo CKD.

**Figure 7:**
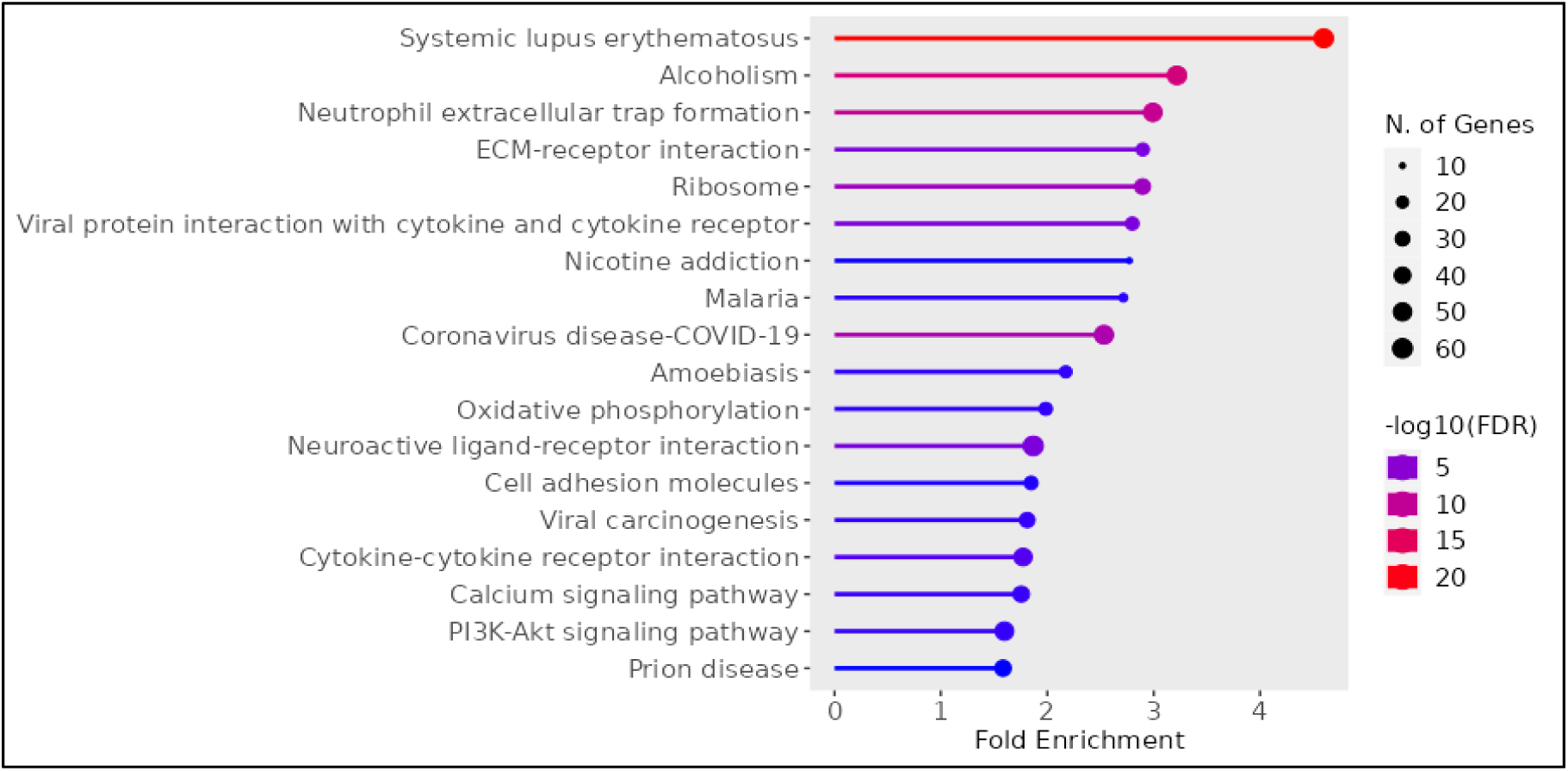
This figure depicts the pathway enrichment analysis results, emphasizing the enriched biological pathways linked to the experimental data. This graphic provides insights into the functional pathways that may be influenced or altered by the conditions under investigation.

**Table 2.**
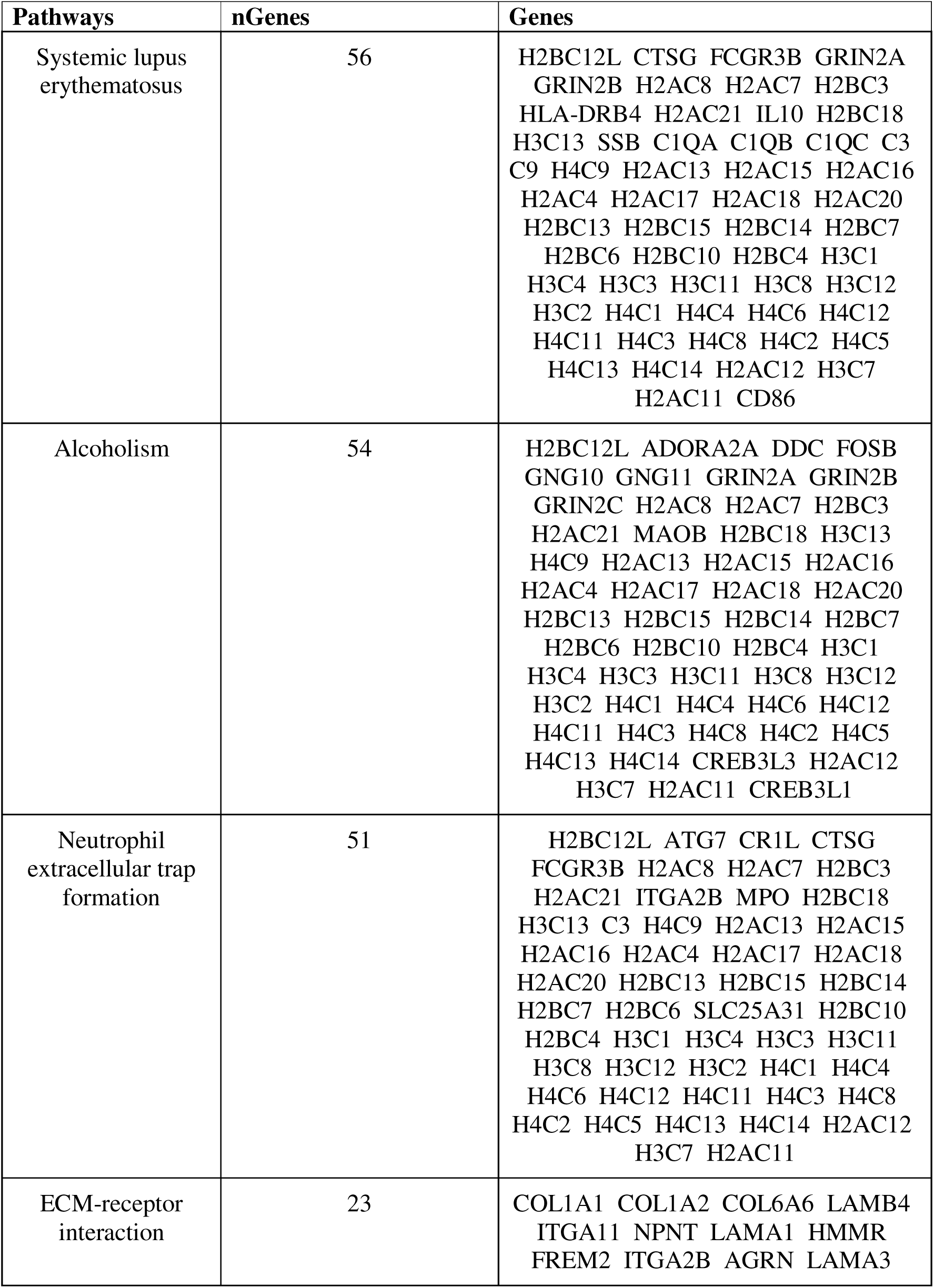
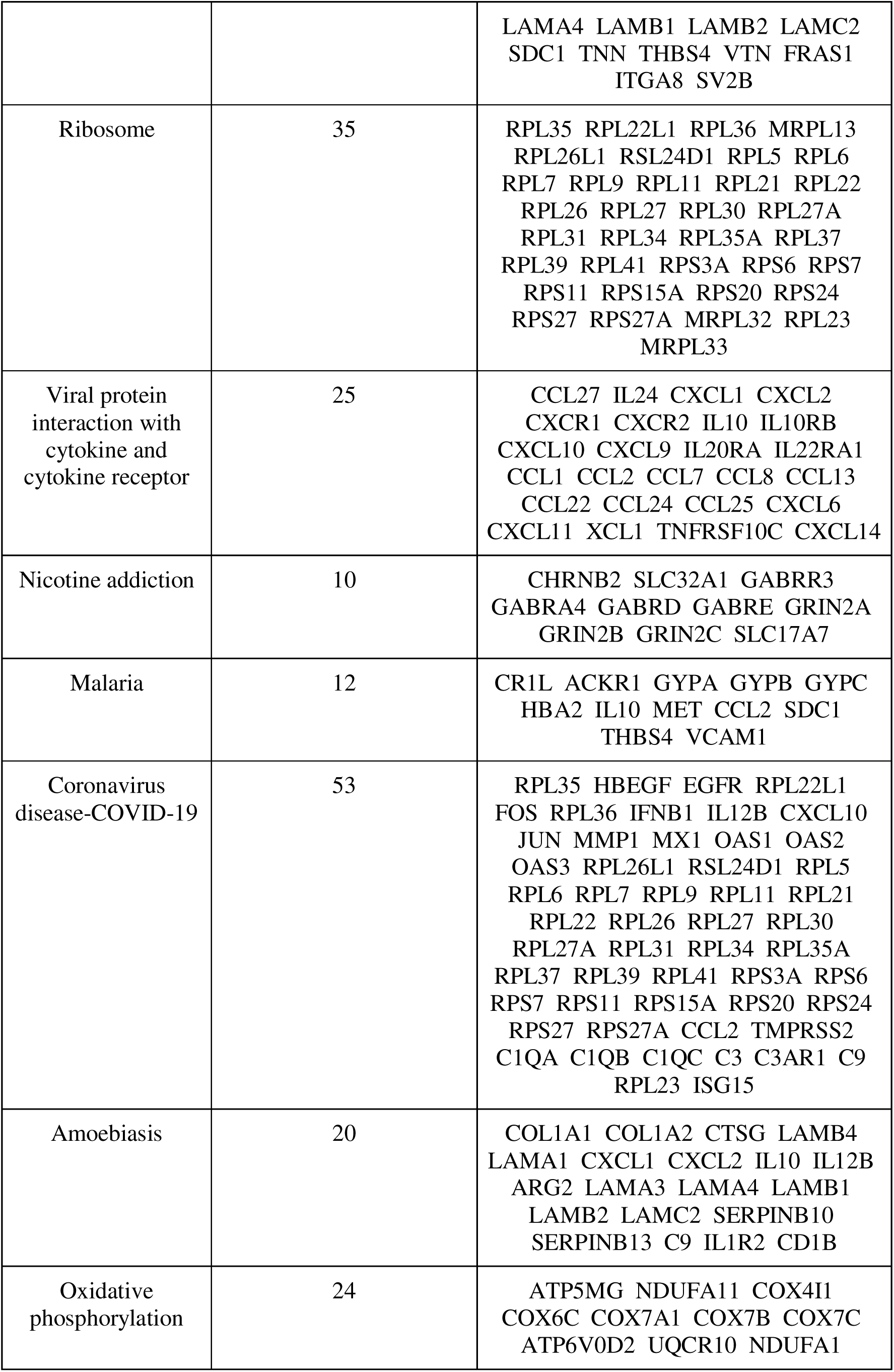
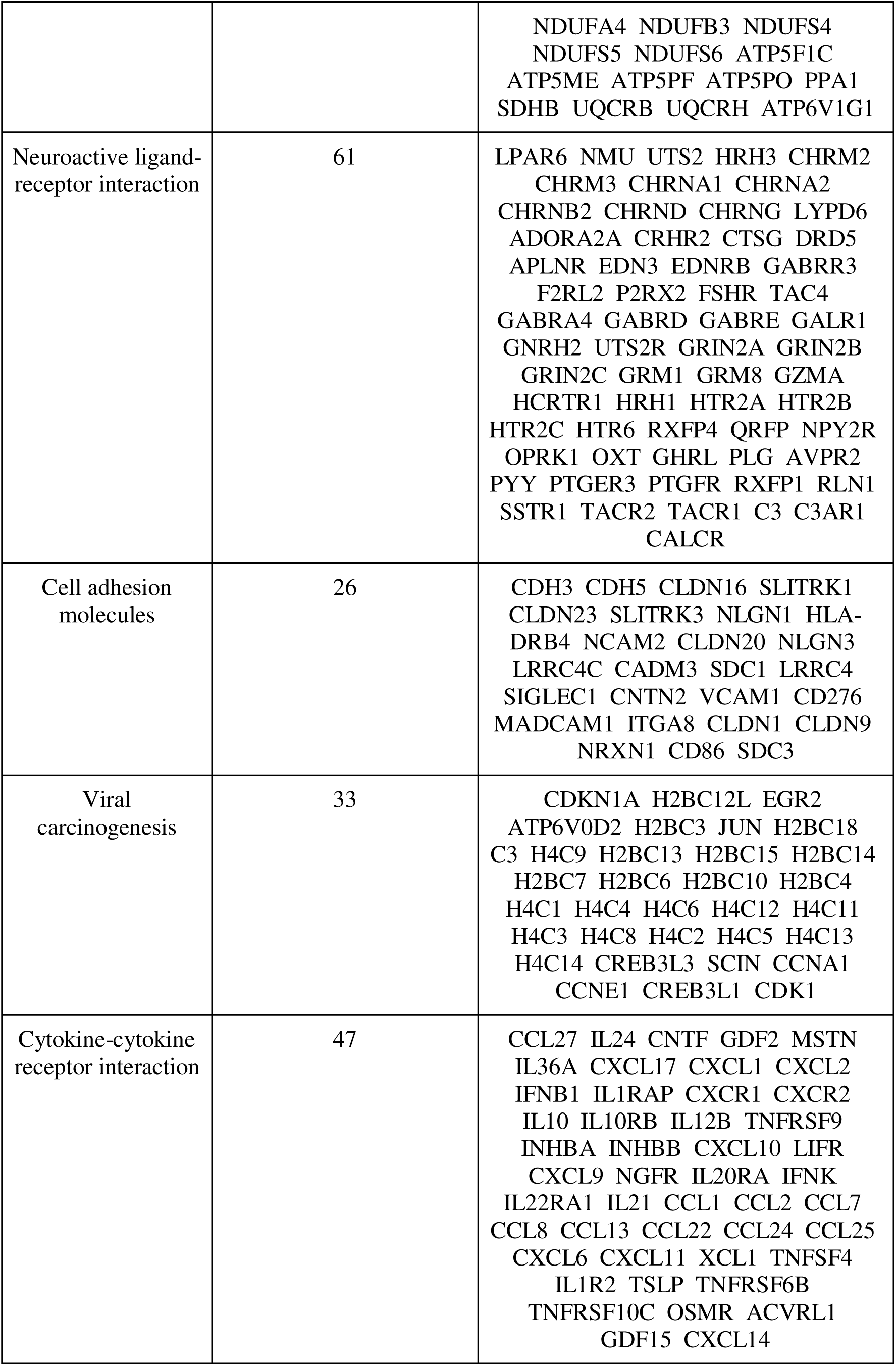
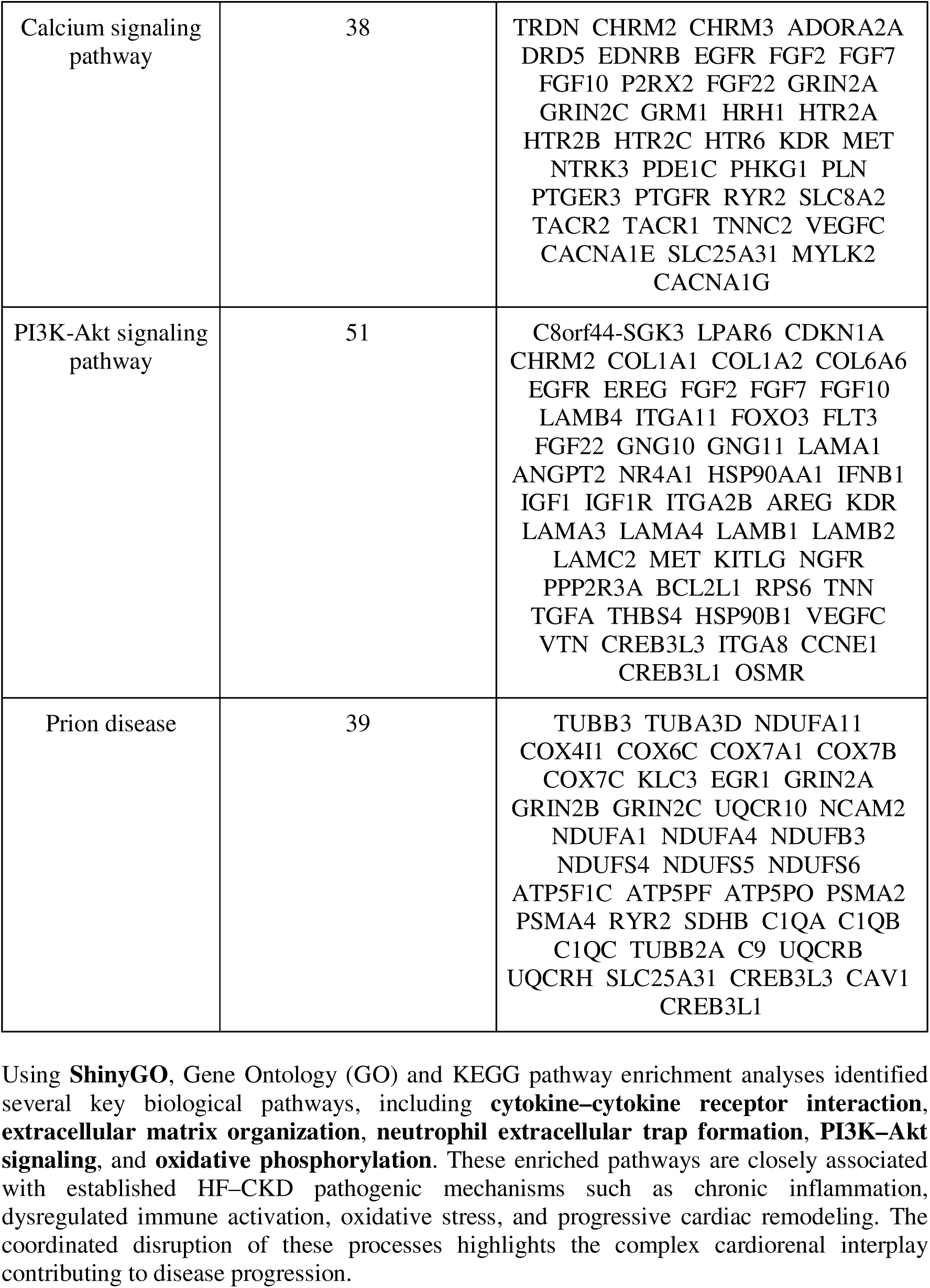
KEGG Pathway Enrichment Analysis for Upregulated Genes Performed Using the ShinyGO Database.

After applying stricter biological cutoffs (log□FC ≥ 5 or ≤ −5), we constructed a reduced, more biologically meaningful PPI network using STRING (confidence ≥ 0.4) and Cytoscape.Key hub genes identified such as **CCL2** (inflammatory chemokine), **ALB** (systemic homeostasis/poor prognosis marker), **EGFR** (fibrosis, hypertrophy signaling), **COL1A2** (extracellular matrix and fibrosis).

## Discussion

Our transcriptomic analysis of PBMCs from CKD patients with HF uncovered a distinct gene expression signature, with four hub genes (EGFR, ALB, CCL2, COL1A2) implicated in cardiac pathology. Notably, these genes have known relevance to HF biology: CCL2 (MCP-1) is a pro-inflammatory chemokine that drives monocyte recruitment and is elevated in heart failure [86] COL1A2 encodes type I collagen, a key component of myocardial fibrosis [87] EGFR mediates epidermal growth factor signaling that influences cardiomyocyte hypertrophy and intercellular coupling [88] and ALB encodes serum albumin, whose depletion (hypoalbuminemia) marks systemic inflammation and portends worse HF outcomes [54,89],genes reflect inflammatory, fibrotic and stress-response pathways that are likely active in CKD-associated HF. Importantly, our findings underscore the value of PBMC transcriptome profiling as a non-invasive window into disease biology. In line with prior reports, we observed that PBMC gene expression mirrors cardiac and systemic inflammatory/oxidative stress in HF patient [90] Thus, circulating immune-cell transcriptomics may serve as a readily accessible biomarker platform for detecting or monitoring HF in high-risk populations, without the need for endomyocardial biopsy.

Future work should validate these candidate biomarkers through targeted experiments. For example, the differential expression of EGFR, ALB, CCL2 and COL1A2 can be confirmed by quantitative RT-PCR in independent cohorts using established analysis software. Functional studies should perturb these genes in relevant models to assess their effects on hypertrophy, fibrosis or inflammation [34]. Additionally, network tools like GeneMANIA can be used to expand the gene network and predict functionally related partners via a guilt-by-association approach guiding further hypothesis generation [69].

Albumin is already used clinically as a prognostic marker in HF [79]. The identified hub genes and PBMC signature could form the basis of blood-based biomarkers for HF risk stratification in dialysis patients. For example, albumin is already used clinically as a prognostic marker in HF [91], illustrating how our transcriptomic signal (ALB expression) might be translated into patient care. In summary, this study provides a strong foundation for developing non-invasive molecular assays to predict and monitor heart failure in MHD patients, addressing a critical need in cardio-renal medicine. To date, no study has jointly analyzed CKD and HF PBMC transcriptomes alongside PPI-validated hub genes, our work advances beyond earlier, more superficial analyses of the same public datasets.

## Conclusion

Our integrative transcriptomic and network-based analysis identified **CCL2** and **ALB** as central hub genes strongly associated with both **heart failure (HF)** and **chronic kidney disease (CKD)**. Evidence from the Comparative Toxicogenomics Database (CTD) (https://ctdbase.org/) further supports their involvement, demonstrating curated associations with key cardiovascular and renal pathologies [80]. **CCL2**, a chemokine critical for monocyte recruitment and inflammatory signaling, has been extensively linked to cardiac remodeling, hypertensive responses, and progression of kidney injury. Likewise, **ALB**, a fundamental serum protein regulating osmotic balance and systemic homeostasis, is significantly associated with adverse cardiovascular outcomes, hypoalbuminemia-related complications, and declining renal function. Their presence across both cardiac and renal disease contexts underscores the interconnected nature of inflammation, metabolic dysregulation, and organ crosstalk in patients with heart failure undergoing maintenance hemodialysis [45,46]. These findings not only validate the biological relevance of our hub genes but also highlight their potential as **biomarker candidates** for early detection, risk stratification, and therapeutic targeting in cardiorenal syndrome [52,60,61].

This study identifies four biologically meaningful hub genes (**CCL2, ALB, EGFR, and COL1A2**) that play central roles in the interconnected pathophysiology of chronic kidney disease and heart failure. The transcriptomic alterations observed in PBMCs reflect key mechanisms such as inflammation, extracellular matrix remodeling, immune dysregulation, and oxidative stress, offering valuable insight into the cardiorenal axis. These findings establish a strong basis for future research aimed at developing **PBMC-based biomarkers** for early detection of heart failure in CKD patients. Additionally, developing machine learning-based predictive models in R (caret, mlr3) [82,83] could enhance diagnostic precision, underscore the translational value of this study and pave the way for personalized approaches to managing heart and kidney diseases.

## Abbreviations

ALB: Albumin
BAM: Binary Alignment Map
CABMR: Chronic Active Antibody-Mediated Rejection
CCL2: Chemokine (C-C motif) ligand 2 (also MCP-1)
CKD: Chronic Kidney Disease
COL1A2: Collagen Type I Alpha 2 Chain
CTD: Comparative Toxicogenomics Database
DEGs: Differentially Expressed Genes
EGFR: Epidermal Growth Factor Receptor
ENA: European Nucleotide Archive
ESRD: End-Stage Renal Disease
FC: Fold Change
FDR: False Discovery Rate
GEO: Gene Expression Omnibus
GO: Gene Ontology
HF: Heart Failure
KEGG: Kyoto Encyclopedia of Genes and Genomes
MCP-1: Monocyte Chemoattractant Protein-1 (synonym for CCL2)
MHD: Maintenance Hemodialysis
OT: Operational Tolerance
PBMCs: Peripheral Blood Mononuclear Cells
PPI: Protein–Protein Interaction
qRT-PCR: Quantitative Reverse Transcription Polymerase Chain Reaction
RNA-seq: RNA Sequencing
SRA: Sequence Read Archive
STRING: Search Tool for the Retrieval of Interacting Genes/Proteins

## Declarations

## Data Availability

All data produced in the present study are available upon reasonable request to the authors

## Acknowledgments

We wish to Acknowledge Dr. Omics Research Labs for their invaluable support and collaboration, which significantly contributed to the success of this research. We would like to express our sincere gratitude for providing the necessary facilities and encouragement to carry out this work.

## Authors Declaration

M.S., M.C., L.P., S.N., and E.S. contributed to Methodology, Literature Review, Data Curation, Formal Analysis, and Visualization. D.P. and S.M. were responsible for Conceptualization, Study Design, Draft Writing, Review & Editing, Supervision, and also contributed to Data Interpretation, Table and Figure Preparation, and Conclusion Drafting.

D.S. provided Supervision, critically Reviewed the Analysis, and contributed to Validation and Final Interpretation.

## Conflicts of interest

The authors declare that they have no conflicts of interest.

## Ethical approval

Not applicable.

## Consent to participate

Not applicable.

## Consent to publication

Not applicable.

## Funding

This research was funded by DrOmics Labs Private Limited India.

## Copyright

© The Author(s) 2025.

## Availability of data and materials

Raw RNA-seq data used in this study are publicly available. Case PBMC transcriptomes from CKD patients with heart failure (HF) are accessible under SRA accession **SRX23265333**, and control PBMC transcriptomes from healthy individuals under **SRX19031772**. FASTQ files were retrieved via the **European Nucleotide Archive (ENA)**: https://www.ebi.ac.uk/ena/browser/.

## Notes

### Competing Interest Statement

The authors have declared no competing interest.

### Funding Statement

This study was funded by Dr.Omics Labs

