## Supplementary Table S2 for "Transcriptomic Profiling of Peripheral Blood Mononuclear Cells Reveals Key Molecular Signatures in Chronic Kidney Disease Patients with Heart Failure"

| **Gene Ontology (GO)** | **No. of genes involved (nGenes)** | **Genes** |
| --- | --- | --- |
| **BIOLOGICAL PROCESS** | | |
| Cytoplasmic Translation | 39 | RPL26L1 ZC3H15 RPL31 RPL6 RWDD1 RPL22 RPL36 ZNF385A RPL26 RPL22L1 RPL9 RPL35A RPS20 EIF3E RPL34 RPL21 RPL5 RPL23 RPL27 RPS15A RPL35 RPS6 RPS24 RPS11 RPL11 RPS27A RPS3A RPL37 RPL7 RPL30 RPL27A RPS7 RPS27 RPS17 EIF3C RPL39 RPL41 RPS18 IGF2BP1 |
| Blood Vessel Development | 88 | ITGA2B LAMA1 RASIP1 COL1A1 AMOTL2 KDR EPHB2 APLNR ANXA1 ACVRL1 VEGFC BMPER COL22A1 SCG2 DCN NCL F3 TNN NR4A1 PPARG KLF4 SULF1 FGF2 WARS1 CXCL10 GDF2 ASB4 ABCC8 FOXC1 NGFR FGF10 FAP LRP2 MMP2 CMA1 CAV1 WNT2 TSPAN12 HOXA13 CCL24 MEOX2 UNC5B LAMA4 PRRX1 PLG EGR2 MMP19 EREG C3 CITED1 TNNI3 APOE TBX3 IL10 PROK1 GPC3 MMP14 TBXT SHH VSTM4 E2F7 TMEM100 ZFPM2 SEMA3E C3AR1 CLEC14A JUN CDH5 ANXA2 GJC1 CCBE1 TAFA5 SAT1 IL12B CREB3L1 HOXA5 WNT5A RRAS COL1A2 MESP1 GRN EGR1 ADAMTS9 ROBO2 GPNMB CCL2 ID1 NPR1 |
| Circulatory System Development | 128 | ITGA2B PKP2 MYLK2 LAMA1 RASIP1 COL1A1 AMOTL2 SOX9 KDR EPHB2 APLNR ANXA1 TBX3 ACVRL1 VEGFC TBXT BMPER MESP1 COL22A1 ZFPM2 SCG2 CDH5 MESP2 NRAP DCN IGF1 FOXC1 NCL F3 TNN NR4A1 PPARG KLF4 SULF1 FGF2 WARS1 CXCL10 GDF2 ASB4 ABCC8 COL11A1 NGFR FGF10 FAP LRP2 MMP2 CMA1 CAV1 WNT2 TSPAN12 HOXA13 CCL24 MEOX2 UNC5B LAMA4 WNT5A PRRX1 PLG EGR2 MMP19 CDKN1A EREG C3 CITED1 ID1 TNNI3 APOE IL10 PROK1 GPC3 FREM2 GJA1 ZIC3 MMP14 ACTC1 LMNA VCAM1 XIRP2 HEYL SHH VSTM4 E2F7 TMEM100 DNAAF3 CDK1 SEMA3E C3AR1 SH3PXD2B CLEC14A IRX3 JUN ANXA2 GJC1 CCBE1 SLIT3 FLRT2 MYH6 ODAD3 RYR2 ARHGEF15 TAFA5 SAT1 ADAP2 IL12B CREB3L1 DSG2 MYH7 HOXA5 CPE TBX18 RRAS NTRK3 EGFR COL1A2 NPY2R GRN ID2 EGR1 OLFM1 RBP4 ADAMTS9 PITX2 ROBO2 CACNA1G GPNMB CCL2 NPR1 CRIP1 |
| Animal Organ Morphogenesis | 116 | LAMA3 LAMC2 FGF10 HOXA9 LAMB4 MYLK2 LAMA1 ATRNL1 TBX3 FGF2 FGF7 ASXL3 IGFN1 TBXT MESP1 LAMB2 ODAPH MESP2 AGRN MYO7A LTF TFAP2A FOXC1 COL11A1 NGFR ITGA8 LRP2 MMP2 RP1 CAV1 WNT2 HOXA5 TSPAN12 HOXA13 COL1A1 AREG WNT5A SDC1 PRRX1 NR5A2 SOX9 PAX1 KDR EPHB2 FST LRP4 APLNR SULF1 FRAS1 LRIG3 ACVRL1 EGFR GPC3 PCDH15 VEGFC MMP16 ZIC3 MMP14 ACTC1 XIRP2 HESX1 HEYL HCN1 SHH OSR2 FAT3 HTRA1 TMEM100 WDR72 NPNT IRX2 SIX2 PKHD1 RXFP1 SCUBE2 IRX3 JUN FJX1 TSKU GPC6 SLIT3 FLRT2 SAPCD2 RYR2 PAX9 RORB MAFB FOXD1 HOXB7 CRIP1 INHBA PKP2 MYH7 CPE TRPV4 SSUH2 TNNI3 PITX2 COL1A2 ZFPM2 NPY2R MYH6 TENM3 MYO3A TBX18 ID2 CITED1 OLFM1 RBP4 ACTG2 ROBO2 DCN TMEM176B CCL2 ONECUT2 EREG |
| Tissue Development | 206 | LAMA3 LAMC2 LAMB4 MYLK2 LAMA1 MEOX2 ATRNL1 COL12A1 TBX18 NR5A2 SOX9 KDR KRT17 ANXA1 TBX3 KRT7 SEMA3D CLEC3B TBXT MESP1 KRT80 COL22A1 SEMA3E KRT8 KRT6C KRT4 KRT74 KRT19 LAMB2 ODAPH KRT10 MESP2 AGRN NRAP KRT6A KRT81 PPARG PITX2 IGF1 FGF10 MET COL1A1 CSTA INHBA FST TFAP2A FGF2 FGF7 SHH CXCL10 UNC5C CES1 SETSIP TUBB3 GDF2 ROS1 KITLG FOXC1 COL11A1 NGFR EYA2 ITGA8 RBFOX1 LRP2 MMP2 HNF4A FERMT1 TIMP1 CASP14 CNFN RASIP1 CAV1 WNT2 HOXA5 HOXA13 AREG HBEGF STC2 WNT5A SDC1 PRRX1 CENPF ONECUT2 EPCAM TNN EGR1 NEUROG3 EGR2 NR4A1 EDN3 SOX21 C3 PAX1 CITED1 ID1 POSTN TPT1 HMGCS2 LRP4 APLNR EHF EDNRB IL10 KLF4 MYO7A SULF1 MSTN FRAS1 LGR5 ACVRL1 PLK4 IL17RD EGFR GPC3 PCDH15 VEGFC FREM2 KLF10 ZIC3 MMP14 HYDIN ACTC1 TCHH LMNA ASGR2 ATF3 XIRP2 CLDN1 HESX1 HEYL BMPER COL1A2 OSR2 TMC1 GJB2 E2F7 TMEM100 WDR72 NPNT VAMP5 ZFPM2 CDK1 FOS IRX2 SIX2 FSHR PKHD1 RXFP1 RPS7 DMRT2 SCUBE2 IRX3 JUN HES7 BGN GJC1 GPC6 STAC3 SAPCD2 ISG15 GJB5 DACT3 MYH6 ARC RYR2 PAX9 MAFB HSBP1 FOXD1 OC90 HOXB7 MMP8 CRIP1 DSG2 PKP2 CDH3 MYH7 MYH14 TRPV4 TNNI3 CHRNA1 GJA1 TPPP3 CDH5 ROBO2 SCIN LTF EPHA3 ESRP1 ID2 PLAAT1 OLFM1 RBP4 RAB13 ACTG2 ADAMTS9 AMER2 SH3PXD2B TSKU CACNA1G CHRD EDF1 EREG PYY EMP1 HLA-DRB1 |
| Anatomical Structure Morphogenesis | 258 | HECW1 ITGA2B EPHA3 LAMA3 FOXC1 LAMC2 FGF10 EPHA8 HOXA9 CDH7 APLP2 LAMB4 MYLK2 LAMA1 TNNT1 RASIP1 ATRNL1 UNC5B COL12A1 TBX18 CDH6 AMOTL2 PLPPR4 SLIRP TNN SLITRK3 NRK SOX21 SOX9 KDR MNX1 EPHB2 ANXA1 TBX3 FGF2 ACVRL1 FGF7 ASXL3 APP FBN3 CNTN4 LRRC4C CDH22 VEGFC SEMA3D UNC5D IGFN1 PITX2 TBXT BMPER MESP1 COL22A1 SEMA3E MAP6 SCG2 LAMB2 ODAPH SLITRK1 CDH5 UNC5C EPHA10 SLIT3 ROBO2 KRT10 DCC MESP2 AGRN NRAP FOXD1 TUBB3 MYO7A NLGN1 DCN LTF FBLN1 RIMS1 OLFM4 MET CCL24 CCL7 CCL2 NCL F3 INHBA NR4A1 PPARG KLF4 TFAP2A SULF1 WARS1 OTX2 CXCL10 KRT8 KRT19 RIMS2 CCL13 GDF2 ASB4 ABCC8 ETV1 COL11A1 NGFR ITGA8 FAP HSP90AA1 LRP2 MMP2 CMA1 TYRO3 PALM HNF4A RP1 MYH14 CAV1 WNT2 HOXA5 TSPAN12 HOXA13 MEOX2 COL1A1 AREG HBEGF WNT5A SF3B6 SDC1 PRRX1 NR5A2 SYF2 STMN1 ONECUT2 NEUROG3 EGR2 MMP19 EDN3 OPN5 EREG C3 PAX1 CITED1 ID1 KRT17 TNNI3 APOE NES FST LRP4 APLNR IL10 RPS6 MYOF CXCL9 FRAS1 LRIG3 LGR5 BARHL2 PROK1 EGFR GPC3 PCDH15 FREM2 PTPRD UCHL1 MMP16 ZIC3 MMP14 IGF2BP1 ACTC1 ZNF385A ELAVL4 XIRP2 HESX1 HEYL HCN1 SHH OSR2 DCSTAMP FAT3 VSTM4 E2F7 HTRA1 TMEM100 WDR72 PRTG NPNT TM4SF1 ZFPM2 IRX2 SIX2 PKHD1 RXFP1 C3AR1 RPS7 DMRT2 SCUBE2 CLEC14A IRX3 JUN ADAMTSL1 HES7 FJX1 TSKU ANXA2 GJC1 GPC6 CCBE1 FLRT2 SAPCD2 SHTN1 GJB5 MYH6 ARC RYR2 PAX9 ARHGEF15 RORB MAFB TAFA5 HSBP1 TRNP1 HOXB7 MMP8 SAT1 CRIP1 FERMT1 IL12B CREB3L1 PKP2 MYH7 CPE TRPV4 CRISPLD1 SSUH2 RRAS FLNC PPFIA2 GJA1 COL1A2 NPY2R SPRY4 S100A10 TENM3 GRN EPB41L3 MYO3A ID2 OLFM1 ANO1 RBP4 ACTG2 ADAMTS9 KRT6A GPNMB MLXIPL ANOS1 EYA2 TMEM176B NPR1 PHLDA3 |
| Animal Organ Development | 309 | MAOB TTBK1 ATP5PF POTEE LAMA3 PKP2 LAMC2 FGF10 HOXA9 LAMB4 MYLK2 LAMA1 RP1 MET TMEM176B ATRNL1 COL1A1 ADGRL2 FLT3 SOX9 KDR MNX1 APLNR ANXA1 TBX3 KRT7 MYO7A SULF1 TACC2 FGF2 ACVRL1 FGF7 ASXL3 ADGRL3 SEMA3D FCER1G IGFN1 HESX1 CLEC3B TBXT MESP1 KRT80 ZFPM2 SEMA3E KRT8 KRT6C KRT4 KRT74 SIX2 FSHR LAMB2 ODAPH DES TEX19 ROBO2 SPRY4 MESP2 AGRN NRAP MAFB KRT6A KRT81 LILRB2 PPARG VCAM1 LTF IGF1 KITLG FOXC1 WNT2 HOXA5 IL12B WNT5A TPO CSTA INHBA IER3IP1 FST LRP4 TFAP2A C1QC SHH UNC5C ANXA2 HLA-DRB1 TUBB3 CALCR ETV1 HMGB3 F7 PTPRU COL11A1 NGFR EML1 ITGA8 RBFOX1 LRP2 ADAMTS2 MMP2 CMA1 TYRO3 LGALS1 HNF4A FERMT1 SLC32A1 TIMP1 MSLN SLC17A7 RETN CASP14 CNFN CAV1 TSPAN12 HOXA13 MEOX2 AREG FRG1 OAS2 STC2 SDC1 PRRX1 RPL22 NR5A2 PLPPR4 SYF2 CENPF ONECUT2 EPCAM TNN EGR1 PLG NEUROG3 EGR2 PRDX4 MMP19 PFDN5 NR4A1 BATF3 EDN3 CDKN1A OPN5 EREG SOX21 PAX1 CITED1 ID1 KRT17 CDO1 TNNI3 TOP2A SLC6A11 RIDA EPHB2 HMGCS2 RSAD2 EDNRB IL10 KLF4 RPS6 MSTN FRAS1 LRIG3 LGR5 SSTR1 NTRK3 APP PLK4 BARHL2 CNTN4 IL17RD CXCL14 EGFR GPC3 CDH22 PCDH15 VEGFC FREM2 GFRA1 GJA1 KLF10 MMP16 ZIC3 MMP14 HYDIN IGF2BP1 ACTC1 TCHH PGLYRP3 LMNA ZNF385A ASGR2 ELAVL4 ATF3 XIRP2 CLDN1 PTX3 HEYL HMGB2 HCN1 BMPER COL1A2 OSR2 DCSTAMP TMC1 PCDH19 FAT3 GJB2 VSTM4 E2F7 HTRA1 TMEM100 WDR72 DNAAF3 NPNT VAMP5 CDK1 FOS IRX2 PKHD1 KRT19 RXFP1 RPS7 COPRS C1QB SH3PXD2B SEZ6L2 GPR149 SCUBE2 IRX3 JUN FJX1 BGN CLEC4G TSKU GJC1 GPC6 CCBE1 SLIT3 FLRT2 STAC3 TAL2 SAPCD2 DCC ISG15 GJB5 DACT3 MYH6 ODAD3 RYR2 PAX9 ARHGEF15 RORB FOXD1 OC90 TRNP1 CLEC5A HOXB7 GDF2 UTF1 ADAP2 KRT10 CRIP1 SCIN DSG2 CDH3 CHRD MYH7 CHRDL1 MYH14 CPE TRPV4 TBX18 STMN1 SSUH2 RRAS RBP4 CHRNA1 TPPP3 PITX2 NDUFS4 NPY2R TENM3 EPHA3 MECOM MYO3A ESRP1 ID2 PLAAT1 OLFM1 NES CTSL MGARP IL21 MDGA2 RAB13 ACTG2 ADAMTS9 MLF1 CACNA1G CD86 GRIN2A CSNK1E GRIN2B DCN CCL2 HBEGF OTX2 CXCL10 IRF7 |
| System Development | 349 | MAOB TTBK1 ATP5PF POTEE HECW1 ITGA2B SCIN ANOS1 STMN4 EPHA3 LAMA3 PKP2 LAMC2 FGF10 EPHA8 RBFOX1 HOXA9 APLP2 TYRO3 MYLK2 LAMA1 RP1 RASIP1 MET WNT2 TMEM176B UNC5B COL1A1 AMOTL2 WNT5A ADGRL2 PLPPR4 STMN1 TNN SLITRK3 FLT3 NEUROG3 NRK SOX9 KDR MNX1 EPHB2 LRP4 APLNR ANXA1 TBX3 SULF1 TACC2 FGF2 ACVRL1 MDGA2 FGF7 NTRK3 APP CNTN4 MANF MAL2 LRRC4C ADGRL3 VEGFC GFRA1 LGI4 SEMA3D MMP16 UNC5D ZIC3 MMP14 CLSTN2 FCER1G IGF2BP1 S100A9 IGFN1 HESX1 HEYL TBXT BMPER SHH OSR2 MESP1 COL22A1 NLGN1 ZFPM2 SEMA3E IRX2 SIX2 FSHR MAP6 SCG2 IRX3 SLITRK1 HES7 CDH5 UNC5C TEX19 GPC6 EPHA10 SLIT3 ROBO2 PCLO C3orf70 DCC MESP2 NRAP BRINP2 MAFB TENM3 TUBB3 LILRB2 MYO7A VCAM1 PITX2 DCN LTF IGF1 KITLG FOXC1 RIMS1 HOXA5 CCL2 IL12B NCL TPO F3 INHBA NR4A1 APOE PPARG IER3IP1 FST KLF4 WARS1 PTPRD C1QC OTX2 CXCL10 RIMS2 ANXA2 SHTN1 HLA-DRB1 GDF2 CALCR ASB4 ABCC8 ETV1 HMGB3 COL11A1 PRDM6 NGFR EML1 ITGA8 FAP HSP90AA1 LRP2 ADAMTS2 MMP2 DNAJB11 CMA1 PALM LGALS1 HNF4A SLC32A1 CHRDL1 TIMP1 SLC17A7 RETN CAV1 TSPAN12 HOXA13 CCL24 MEOX2 AREG TRPV4 LAMA4 STC2 SDC1 PRRX1 RPL22 ONECUT2 EPCAM KIAA1217 EGR1 PLG EGR2 PRDX4 MMP19 PFDN5 BATF3 EDN3 CDKN1A OPN5 EREG C3 PAX1 CITED1 ID1 RRAS CHAC1 TNNI3 OLFM1 C1QL1 TOP2A SLC6A11 RIDA SNAP25 NES BEX1 HMGCS2 RSAD2 EDNRB SCN2A IL10 RPS6 PCDH10 FRAS1 LGR5 SSTR1 PLK4 BARHL2 PROK1 S100A8 EGFR GPC3 CDH22 PCDH15 FREM2 GJA1 UCHL1 NCAM2 KLF10 HYDIN ACTC1 PGLYRP3 LMNA ZNF385A ELAVL4 XIRP2 CLDN1 PTX3 HMGB2 HCN1 DCSTAMP TMC1 PCDH19 FAT3 GJB2 VSTM4 E2F7 HTRA1 TMEM100 PRTG DNAAF3 NPNT CDK1 FOS PKHD1 KRT19 RXFP1 C3AR1 RPS7 LAMB2 HOXC5 DMRT2 C1QA EXO1 SH3PXD2B SEZ6L2 GPR149 LSM1 SCUBE2 NR2F1 CLEC14A JUN ADAMTSL1 FJX1 BGN CLEC4G TSKU GJC1 CCBE1 GRIN2A FLRT2 TAL2 ISG15 HES4 VWC2 GJB5 MYH6 S100A10 ODAD3 ARC RYR2 ARHGEF15 RORB TAFA5 FOXD1 TRNP1 CLEC5A HOXB7 SAT1 TFAP2A UTF1 ADAP2 CRIP1 CREB3L1 KRT8 DSG2 CHRD MYH7 CPE TBX18 CENPF COX7B RBP4 PPFIA2 RAB13 TPPP3 NDUFS4 COL1A2 NPY2R GRN EPB41L3 MECOM ESRP1 SEPTIN4 ID2 PLAAT1 ANO1 CTSL MGARP IL21 SCARB2 ADAMTS9 MLF1 AGRN CACNA1G CD86 CHST8 GPNMB CSNK1E GRIN2B PYY CHRM3 BTG4 TAGLN3 NPR1 CHRM2 NTM IRF7 LSAMP |
| Cellular Developmental processes | 332 | HSPE1 HECW1 ETV1 ANOS1 STMN4 EPHA3 LAMA3 FOXC1 LAMC2 EYA2 FGF10 EPHA8 APLP2 LAMB4 HNF4A LAMA1 RP1 TNNT1 CAV1 MET WNT2 TMEM176B ATRNL1 UNC5B COL12A1 TBX18 WNT5A ID2 PLPPR4 STMN1 SLIRP TNN SLITRK3 FLT3 NEUROG3 NRK SOX21 SOX9 CITED1 ID1 KDR KRT17 MNX1 PPARG EPHB2 FST LRP4 ANXA1 TBX3 EHF KRT7 MYOF FGF2 FGF7 NTRK3 APP CNTN4 MANF EGFR POMZP3 LRRC4C LGI4 SEMA3D UNC5D FCER1G CCDC42 S100A9 HEYL TBXT SHH SOHLH1 KRT80 COL22A1 ZFPM2 SEMA3E KRT8 KRT6C KRT4 KRT74 IRX2 KRT19 MAP6 LAMB2 NR2F1 FAM9B IRX3 SLITRK1 HES7 UNC5C EPHA10 SLIT3 ROBO2 KRT10 DCC NRAP BRINP2 MAFB KRT6A KRT81 TENM3 FOXD1 ETV3L TUBB3 LILRB2 MYO7A TTBK1 VCAM1 PITX2 NLGN1 LTF IGF1 GRN FBLN1 RIMS1 MYLK2 OLFM4 CEACAM5 HOXA5 CCL2 COL1A1 IL12B CSTA INHBA EREG APOE TFAP2A MSTN IL21 C1QC OTX2 ZNF503 RIMS2 ANXA2 FFAR4 SHTN1 HLA-DRB1 CES1 SETSIP EID1 CALCR ASB4 ABCC8 HMGB3 MSR1 ROS1 KITLG PTPRU COL11A1 PRDM6 NGFR EML1 TRIP13 ITGA8 RBFOX1 HOXA9 HSP90AA1 LRP2 MECOM MMP2 DNAJB11 TYRO3 LGALS1 CHRDL1 HTR2A MEDAG RETN CASP14 CNFN RASIP1 CFAP69 HOXA13 EDF1 SEPTIN4 AREG DDX25 TRPV4 LAMA4 SDC1 PRRX1 RPL22 NR5A2 CENPF ONECUT2 EGR1 PLG EGR2 MMP19 NR4A1 BATF3 EDN3 C3 PAX1 RRAS FLNC CHAC1 TNNI3 C1QL1 TOP2A SNAP25 NES BEX1 TEX15 RSAD2 APLNR EDNRB KLF4 RPS6 SULF1 BRDT CXCL9 LGR5 ACVRL1 PLK4 BARHL2 S100A8 IL17RD RPS3A CXCL14 HTR2C GPC3 TLCD3B PCDH15 VEGFC SLC25A31 GFRA1 PTPRD UCHL1 NCAM2 KLF10 ZIC3 MMP14 HYDIN IGF2BP1 ACTC1 TCHH PGLYRP3 LMNA ZNF385A ELAVL4 ATF3 CFAP221 CLDN1 ADAMTS9 PTX3 HESX1 HMGB2 HCN1 OSR2 DCSTAMP TMC1 FAT3 E2F7 HTRA1 TMEM100 PRTG MESP1 NPNT VAMP5 CXCL10 CDK1 FABP4 FOS SIX2 FSHR RXFP1 RPS7 C1QA SH3PXD2B LARP7 GPR149 LSM1 SCUBE2 JUN ADAMTSL1 MLF1 CFAP65 TEX19 CLEC4G TSKU GJC1 GRIN2A CFAP91 FLRT2 STAC3 ISG15 MORN2 AGRN HES4 VWC2 GJB5 DACT3 MYH6 S100A10 ARC SYCP1 RORB HSBP1 CLEC5A HOXB7 GDF2 MMP8 IL10 SCIN LMO3 PKP2 CDH3 CHRD MYH14 EPCAM PPFIA2 RAB13 GJA1 CDH5 SPRY4 EPB41L3 ESRP1 PLAAT1 OLFM1 ANO1 CTSL MGARP SCARB2 MDGA2 ADGRL3 CREB3L1 PKHD1 CD86 CSNK1E PYY BTG4 NTM IRF7 |
| Cell Differentiation | 330 | HSPE1 HECW1 ETV1 ANOS1 STMN4 EPHA3 LAMA3 FOXC1 LAMC2 EYA2 FGF10 EPHA8 APLP2 LAMB4 HNF4A LAMA1 RP1 TNNT1 CAV1 MET WNT2 TMEM176B ATRNL1 UNC5B COL12A1 TBX18 WNT5A ID2 PLPPR4 STMN1 TNN SLITRK3 FLT3 NEUROG3 NRK SOX21 SOX9 CITED1 ID1 KDR KRT17 MNX1 PPARG EPHB2 FST LRP4 ANXA1 TBX3 EHF KRT7 MYOF FGF2 FGF7 NTRK3 APP CNTN4 MANF EGFR POMZP3 LRRC4C LGI4 SEMA3D UNC5D FCER1G CCDC42 S100A9 HEYL TBXT SHH SOHLH1 KRT80 COL22A1 ZFPM2 SEMA3E KRT8 KRT6C KRT4 KRT74 IRX2 KRT19 MAP6 LAMB2 NR2F1 FAM9B IRX3 SLITRK1 HES7 UNC5C EPHA10 SLIT3 ROBO2 KRT10 DCC NRAP BRINP2 MAFB KRT6A KRT81 TENM3 FOXD1 ETV3L TUBB3 LILRB2 MYO7A TTBK1 VCAM1 PITX2 NLGN1 LTF IGF1 GRN FBLN1 RIMS1 MYLK2 OLFM4 CEACAM5 HOXA5 CCL2 COL1A1 IL12B CSTA INHBA EREG APOE TFAP2A MSTN IL21 C1QC OTX2 ZNF503 RIMS2 ANXA2 FFAR4 SHTN1 HLA-DRB1 CES1 SETSIP EID1 CALCR ASB4 ABCC8 HMGB3 MSR1 ROS1 KITLG PTPRU COL11A1 PRDM6 NGFR EML1 TRIP13 ITGA8 RBFOX1 HOXA9 HSP90AA1 LRP2 MECOM MMP2 DNAJB11 TYRO3 LGALS1 CHRDL1 HTR2A MEDAG RETN CASP14 CNFN RASIP1 CFAP69 HOXA13 EDF1 SEPTIN4 AREG DDX25 TRPV4 LAMA4 SDC1 PRRX1 RPL22 NR5A2 CENPF ONECUT2 SLIRP EGR1 PLG EGR2 MMP19 NR4A1 BATF3 EDN3 C3 PAX1 RRAS FLNC CHAC1 TNNI3 C1QL1 TOP2A SNAP25 BEX1 TEX15 RSAD2 APLNR EDNRB KLF4 RPS6 SULF1 BRDT CXCL9 LGR5 ACVRL1 PLK4 BARHL2 S100A8 IL17RD RPS3A CXCL14 HTR2C GPC3 TLCD3B PCDH15 VEGFC SLC25A31 GFRA1 PTPRD UCHL1 NCAM2 KLF10 ZIC3 MMP14 HYDIN IGF2BP1 ACTC1 TCHH PGLYRP3 LMNA ZNF385A ELAVL4 ATF3 CFAP221 CLDN1 ADAMTS9 PTX3 HESX1 HMGB2 HCN1 OSR2 DCSTAMP TMC1 FAT3 E2F7 HTRA1 TMEM100 PRTG MESP1 NPNT VAMP5 CXCL10 CDK1 FABP4 FOS SIX2 FSHR RXFP1 RPS7 C1QA SH3PXD2B LARP7 GPR149 LSM1 SCUBE2 JUN ADAMTSL1 MLF1 CFAP65 TEX19 CLEC4G TSKU GJC1 GRIN2A CFAP91 FLRT2 STAC3 ISG15 MORN2 AGRN HES4 VWC2 GJB5 DACT3 MYH6 S100A10 ARC SYCP1 RORB HSBP1 CLEC5A HOXB7 GDF2 MMP8 IL10 SCIN LMO3 PKP2 CDH3 CHRD EPCAM PPFIA2 RAB13 GJA1 CDH5 SPRY4 EPB41L3 ESRP1 PLAAT1 OLFM1 ANO1 CTSL MGARP SCARB2 MDGA2 ADGRL3 CREB3L1 PKHD1 CD86 CSNK1E PYY BTG4 NTM IRF7 |
| **CELLULAR COMPONENTS** | | |
| Cytosolic Large Ribosomal Subunit | 22 | RPL31 RPL6 RPL34 RPL22 RPL21 RPL5 RPL23 RPL36 RPL27 RPL35 RPL11 RPL37 RPL7 RPL30 RPL26 RPL9 RPL27A RPL35A RPL39 RPL41 RPL26L1 RPL39L |
| Cytosolic Ribosome | 35 | RPS20 RPL31 RPL6 RPL34 RPL22 RPL21 RPL5 RPL23 RPL36 RPL27 RPS15A RPL35 RPS6 RPS24 RPS11 RPL11 RPS27A RPS3A RPL37 RPL7 RPL30 RPL26 RPL9 RPL27A RPS7 RPS27 RPS17 RPL35A RPL39 RPL41 RPS18 RPL26L1 RPL39L ISG15 HBA2 |
| Ribosomal Subunit | 40 | RPS20 RPL31 RPL6 RPL34 RPL22 RPL21 RPL5 RPL23 RPL36 RPL27 RPS15A RPL35 RPS6 RPS24 RPS11 RPL11 RPS27A RPS3A RPL37 RPL7 RPL30 RPL26 RPL9 RPL27A RPS7 RPS27 RPS17 RPL35A RPL39 RPL41 RPS18 RPL26L1 MRPL32 MRPL47 RPL39L MRPL13 ISG15 CHCHD1 MRPL33 HBA2 |
| Collagen-Containing Extracellular Matrix | 71 | DCN LAMA3 ITIH1 F7 LAMC2 COL11A1 FBLN1 FCN1 MMP2 CMA1 LGALS1 CTSG LAMA1 MXRA5 LGALS3BP COL1A1 COL12A1 LAMA4 WNT5A MMP8 PLG COL21A1 APOE POSTN ANXA1 CTSL SULF1 LUM TINAGL1 S100A8 LEFTY2 HMCN2 SERPING1 C1QC SDC3 S100A9 CLEC3B ANXA5 SHH COL1A2 HTRA1 HSP90B1 NPNT LAMB2 C1QA CLEC14A VWA1 BGN ANXA2 PRG2 THSD4 AGRN S100A10 LAMB4 ATRNL1 TNN FGF10 WNT2 F3 ZG16 HSP90AA1 ADAMTS2 TIMP1 FRAS1 FBN3 GPC3 FREM2 LAD1 GPC6 VWC2 ADAMTS9 |
| Ribosome | 44 | RPS20 RPL31 RPL6 RPL34 RPL22 RPL21 RPL5 RPL23 RPL36 RPL27 RPS15A RPL35 RPS6 RPS24 RPS11 RPL11 RPS27A RPS3A RPL37 RPL7 RPL30 RPL26 RPL9 RPL27A RPS7 RPS27 RPS17 RPL35A RPL39 RPL41 RPS18 RPL26L1 MRPL32 MRPL47 RPL39L MRPL13 ISG15 CHCHD1 MRPL33 OAS1 HBA2 RSL24D1 ELAVL4 RPL22L1 |
| Extracellular Matrix | 93 | DCN LAMA3 ITIH1 F7 LAMC2 COL11A1 FBLN1 FCN1 MMP2 CMA1 LGALS1 CTSG LAMA1 MXRA5 LGALS3BP COL1A1 COL12A1 LAMA4 WNT5A MMP8 PLG COL21A1 APOE POSTN ANXA1 CTSL SULF1 LUM TINAGL1 S100A8 LEFTY2 HMCN2 SERPING1 C1QC SDC3 S100A9 CLEC3B ANXA5 SHH COL1A2 HTRA1 HSP90B1 NPNT LAMB2 C1QA CLEC14A VWA1 BGN ANXA2 PRG2 THSD4 AGRN S100A10 ADAMTS2 LAMB4 TIMP1 ATRNL1 TNN LRIG3 FBN3 POMZP3 ZAN COLEC12 ADAMTS9 BMPER COL22A1 LRRC3B OTOG ELFN1 OC90 FGF10 WNT2 F3 ZG16 CCBE1 SPOCK3 HSP90AA1 MMP19 FRAS1 GPC3 FREM2 MMP16 MMP14 LAD1 OLFML2B PTX3 GPC6 OLFML2A VWC2 MMP1 CRISP3 FLRT2 ANOS1 |
| External Encapsulating Structure | 93 | DCN LAMA3 ITIH1 F7 LAMC2 COL11A1 FBLN1 FCN1 MMP2 CMA1 LGALS1 CTSG LAMA1 MXRA5 LGALS3BP COL1A1 COL12A1 LAMA4 WNT5A MMP8 PLG COL21A1 APOE POSTN ANXA1 CTSL SULF1 LUM TINAGL1 S100A8 LEFTY2 HMCN2 SERPING1 C1QC SDC3 S100A9 CLEC3B ANXA5 SHH COL1A2 HTRA1 HSP90B1 NPNT LAMB2 C1QA CLEC14A VWA1 BGN ANXA2 PRG2 THSD4 AGRN S100A10 ADAMTS2 LAMB4 TIMP1 ATRNL1 TNN LRIG3 FBN3 POMZP3 ZAN COLEC12 ADAMTS9 BMPER COL22A1 LRRC3B OTOG ELFN1 OC90 FGF10 WNT2 F3 ZG16 CCBE1 SPOCK3 HSP90AA1 MMP19 FRAS1 GPC3 FREM2 MMP16 MMP14 LAD1 OLFML2B PTX3 GPC6 OLFML2A VWC2 MMP1 CRISP3 FLRT2 ANOS1 |
| Cell Surface | 114 | ITGA2B DSG2 CD209 MET CD86 F3 CD1B ANOS1 SLAMF7 MSR1 NGFR ITGA8 CEACAM6 MSLN CD276 EBI3 CEACAM5 CD69 IL12B LIFR SDC1 CD207 IGLL1 BST2 ULBP2 SULF1 ASGR1 APP GPC3 GFRA1 CLSTN2 FCER1G ASGR2 SDC3 SLAMF9 DCSTAMP FOLR2 PDIA3 GP2 NLGN1 ZPLD1 SCUBE2 CLEC14A CD163 CD163L1 CLEC4G IGSF5 GPC6 DCC CD200R1L CEACAM20 FCN1 LTF FGF10 TYRO3 LIPG WNT2 AREG HBEGF PLG KCNA5 LILRB2 LRP4 ANXA1 CHRNA1 ACVRL1 EGFR PLA2R1 VCAM1 FCRL4 VAMP5 KRT4 CDH5 BGN ANXA2 ROBO2 PTPRT HLA-DRB1 ROS1 HMMR LAMP3 FAP HSP90AA1 LRP2 SLC32A1 PKD2L1 TRPV4 WNT5A TPO EPCAM TNN SLITRK3 C3 TSPAN8 KDR EPHB2 CXCL9 PPFIA2 MMP16 UNC5D ADAMTS9 ANXA5 SHH TMC1 CXCL10 FSHR UNC5C GRIN2A TCN2 KRT10 CLEC5A MRC1 GRIN2B PKHD1 |
| Extracellular Space | 314 | LAP3 MPO ITGA2B GGCT SCIN RPS20 DCN LTF GPRC5A GRN RPL26L1 PSMA4 DSG2 LAMA3 ITIH1 ATP1B3 RPL31 TRHDE FBLN1 HSP90AA1 LRP2 APLP2 DNAJA1 CHMP5 MMP2 F11 LYZ CRISP3 LGALS1 CTSG BPI MXRA5 TIMP1 OLFM4 EIF3E RETN PDCD5 MYH14 CEACAM5 CLEC11A PON3 CPVL PSMA2 BLVRA PTGR1 ALDH3A1 LGALS3BP COL1A1 CPE RPL34 OAS3 COL12A1 LAMA4 MEP1A SUB1 LIFR C9 STC2 CD86 NCL IGFBP2 HSPE1 TPO SDC1 RPL22 STMN1 VAMP8 NPC2 EPCAM CRISPLD1 PLG RPL5 PRDX4 RPL23 C3 SNRPD2 RRAS CSN1S1 TSPAN8 RNASE1 APOE BST2 FBP2 RPL27 ANO1 DDC JCHAIN RIDA TPT1 RPS15A DSG3 ANXA1 CTSL GOLM1 KRT7 ATP6V1G1 PI15 GGH SULF1 UACA MYOF RBP4 SCARB2 PPFIA2 LRIG3 LUM WARS1 APP RPS11 RPL11 TINAGL1 CRNN RAB13 S100A8 RPS27A RPS3A ATP6V0D2 DPYS MAL2 LCN2 LRRC4C SERPING1 TIMM8B FREM2 GFRA1 PTPRR PTPRD DBI RPL30 CABP1 LAD1 C1QC ACTC1 RPL26 VCAM1 ACTG2 S100A9 ALB CLEC3B ANXA5 ENPP6 COL1A2 ALDH1A1 TMEM52B HTRA1 PRTG HSP90B1 PDIA3 LPO SCN11A NPNT VAMP5 RAB3B GP2 RNASE2 HINT1 CDK1 FABP4 KRT8 KRT6C KRT74 PKHD1 KRT19 LAMB2 DES RIMS2 ZNF114 C5orf46 VWA1 PPA1 BGN ANXA2 RPL35A TMPRSS2 ROBO2 FLRT2 KRT10 PCLO PRG2 THSD4 AGRN POTEE HBA2 HLA-DRB1 HBG2 SERPINB13 S100A10 RASSF9 CD177 KRT6A KRT81 HBD RPS18 TXNDC5 PGA5 HP TUBB3 GDF2 ARHGAP23 UTS2 F7 COL11A1 FGF10 CXCL2 FCN1 CPXM1 CHGB CMA1 LIPG CCL22 WFDC1 WNT2 CCL24 CCL7 CCL2 CCL8 CCL1 AREG HBEGF IL12B WNT5A TNN INHBA MMP19 EDN3 COL21A1 EREG IGLL1 APOC1 OLFM1 ULBP2 C1QL1 PYY CCL25 POSTN FST IL36A MSTN FGF2 CXCL9 FGF7 XCL1 LEFTY2 MANF ZAN HMCN2 VEGFC LGI4 SEMA3D DKK2 COLEC12 CD1B OLFML2B S100A12 PTX3 SCGB3A2 BMPER SHH IGFBP6 CXCL10 CXCL11 RNASE6 COL22A1 ZPLD1 SEMA3E SCG2 ZG16 SCUBE2 FJX1 LRRC3B CCL13 TSKU TCN2 OLFML2A OTOG VWC2 TAFA5 ELFN1 SERPINB10 RNASE4 CCL5 IGF1 C1QA FAP CEACAM6 LAMA1 F3 MMP8 CSTA LILRB2 IL10 PCDH15 HMGB2 MUC13 CCBE1 SLIT3 SPOCK3 RPL39 MSR1 KITLG LAMC2 DNAJB11 MSLN EBI3 IL21 TSLP CXCL14 ADAMTS9 GPC6 MMP14 APOL4 CHRD EGFR ANOS1 |
| Extracellular Region | 379 | LAP3 MPO ITGA2B GGCT SCIN RPS20 DCN LTF GPRC5A GRN RPL26L1 PSMA4 DSG2 LAMA3 ITIH1 ATP1B3 RPL31 TRHDE FBLN1 HSP90AA1 LRP2 APLP2 DNAJA1 CHMP5 MMP2 F11 LYZ CMA1 CRISP3 LGALS1 CTSG BPI MXRA5 TIMP1 OLFM4 EIF3E RETN PDCD5 MYH14 CEACAM5 CLEC11A PON3 CPVL PSMA2 BLVRA PTGR1 ALDH3A1 LGALS3BP COL1A1 CPE RPL34 OAS3 COL12A1 LAMA4 MEP1A SUB1 LIFR C9 STC2 CD86 NCL IGFBP2 HSPE1 TPO SDC1 RPL22 STMN1 VAMP8 NPC2 EPCAM CRISPLD1 PLG RPL5 PRDX4 RPL23 C3 SNRPD2 RRAS CSN1S1 TSPAN8 RNASE1 APOE BST2 FBP2 RPL27 ANO1 DDC JCHAIN RIDA TPT1 RPS15A DSG3 ANXA1 CTSL GOLM1 KRT7 ATP6V1G1 PI15 GGH SULF1 UACA MYOF RBP4 SCARB2 PPFIA2 LRIG3 LUM WARS1 APP RPS11 RPL11 TINAGL1 CRNN RAB13 S100A8 RPS27A RPS3A ATP6V0D2 DPYS MAL2 LCN2 LRRC4C SERPING1 TIMM8B FREM2 GFRA1 PTPRR PTPRD DBI RPL30 CABP1 LAD1 C1QC ACTC1 RPL26 VCAM1 ACTG2 S100A9 ALB CLEC3B ANXA5 ENPP6 COL1A2 ALDH1A1 TMEM52B HTRA1 PRTG HSP90B1 PDIA3 LPO SCN11A NPNT VAMP5 RAB3B GP2 RNASE2 HINT1 CDK1 FABP4 KRT8 KRT6C KRT74 PKHD1 KRT19 LAMB2 DES RIMS2 ZNF114 C5orf46 VWA1 PPA1 BGN ANXA2 RPL35A TMPRSS2 ROBO2 FLRT2 KRT10 PCLO PRG2 THSD4 AGRN POTEE HBA2 HLA-DRB1 HBG2 SERPINB13 S100A10 RASSF9 CD177 KRT6A KRT81 HBD RPS18 TXNDC5 PGA5 HP TUBB3 GDF2 ARHGAP23 UTS2 F7 COL11A1 FGF10 CXCL2 FCN1 CPXM1 CHGB LIPG CCL22 WFDC1 WNT2 CCL24 CCL7 CCL2 CCL8 CCL1 AREG HBEGF IL12B WNT5A TNN INHBA MMP19 EDN3 COL21A1 EREG IGLL1 APOC1 OLFM1 ULBP2 C1QL1 PYY CCL25 POSTN FST IL36A MSTN FGF2 CXCL9 FGF7 XCL1 LEFTY2 MANF ZAN HMCN2 VEGFC LGI4 SEMA3D DKK2 COLEC12 CD1B OLFML2B S100A12 PTX3 SCGB3A2 BMPER SHH IGFBP6 CXCL10 CXCL11 RNASE6 COL22A1 ZPLD1 SEMA3E SCG2 ZG16 SCUBE2 FJX1 LRRC3B CCL13 TSKU TCN2 OLFML2A OTOG VWC2 TAFA5 ELFN1 SERPINB10 RNASE4 CCL5 IGF1 C1QA FAP CEACAM6 TPX2 LAMA1 F3 MMP8 CSTA PTGFR LILRB2 IL10 CENPE TSLP PCDH15 HMGB2 MUC13 CCBE1 SLIT3 ISG15 SPOCK3 AKR1B10 RPL39 ANOS1 MSR1 EPHA3 KITLG LAMC2 ADAMTS2 SIGLEC1 OAS1 DNAJB11 CHRD CD209 LAMB4 APOL4 CHRDL1 MSLN PDGFRL EBI3 MET NMU C1orf54 CSTL1 KDR PRRG1 DSC3 KIF23 CLCA2 CEP55 IL21 SPACA3 ASGR1 FBN3 PROK1 CNTN4 GZMA CXCL14 EGFR PLA2R1 MMP16 PGLYRP3 CFAP221 ADAMTS9 FNDC1 FOLR2 VSTM4 NLGN1 MUC3A HTRA3 C1QB RNASE11 ODAPH CD163 CD163L1 ADAMTSL1 SLITRK1 GPC6 EPHA10 LYPD6 MMP1 ARC BRINP2 IFI30 SMIM20 OC90 KIR3DX1 MMP14 NGFR EPHB2 MDGA2 SRP14 LYPD5 LY6E NTM LSAMP |
| **MOLECULAR FUNCTIONS** | | |
| Ubiquitin Ligase Inhibitor Activity | 6 | RPS20 RPL5 RPL23 RPL37 RPS7 RPL11 |
| Chemokine Activity | 15 | CXCL2 CCL22 CCL24 CCL7 CCL2 CCL8 CCL1 CCL25 CXCL9 XCL1 CXCL10 CXCL11 CCL13 CCL5 CXCL14 |
| Structural Constituent of Ribosome | 40 | RPS20 RPS15A RPS6 RPS24 RPS11 RPS27A RPS3A RPS7 RPS17 RPS18 RPL26L1 RPL31 RPL6 MRPL32 RPL22 RPL21 RPL5 RPL23 RPL36 RPL27 MRPL47 RPL35 RPL11 RPL7 RPL30 RPL26 RPL22L1 RPL9 RPL27A MRPL13 RPS27 RPL35A ISG15 RPL34 RPL37 RPL39 RSL24D1 RPL39L RPL41 MRPL33 |
| Extracellular Matrix Structural Constituent | 32 | MXRA5 COL1A1 COL1A2 PRG2 LAMA3 COL11A1 LAMB4 LAMA1 FBN3 GP2 COL22A1 ZPLD1 LAMB2 FBLN1 DCN POMZP3 LAMA4 FRAS1 LUM BGN COL12A1 TINAGL1 MUC3A LAMC2 COL21A1 POSTN HMCN2 NPNT VWA1 THSD4 AGRN ANOS1 |
| Glycosaminoglycan Binding | 38 | HBEGF SULF1 SLIT3 MPO LTF ITIH1 FGF10 CTSG PDCD5 APOE JCHAIN GPNMB PGLYRP3 CLEC3B ANOS1 DCN LAMC2 COL11A1 HMMR APLP2 F11 LIPG CCL7 CCL8 RPL22 POSTN MSTN FGF2 FGF7 APP SHH ZG16 BGN PRG2 SPOCK3 CXCL10 CXCL11 AGRN |
| Heparin Binding | 27 | HBEGF SLIT3 MPO LTF FGF10 CTSG PDCD5 APOE GPNMB CLEC3B ANOS1 LAMC2 COL11A1 APLP2 F11 LIPG CCL7 CCL8 RPL22 POSTN MSTN FGF2 FGF7 APP PRG2 CXCL10 CXCL11 |
| Structural Molecule Activity | 104 | RPS20 MXRA5 COL1A1 RPS15A RPS6 RPS24 RPS11 RPS27A RPS3A COL1A2 RPS7 RPS17 PRG2 RPS18 RPL26L1 LAMA3 COL11A1 RPL31 RPL6 LAMB4 LAMA1 MRPL32 RPL22 RPL21 RPL5 RPL23 RPL36 RPL27 KRT7 MRPL47 RPL35 FBN3 RPL11 RPL7 MAL2 RPL30 RPL26 RPL22L1 RPL9 RPL27A KRT80 GP2 COL22A1 ZPLD1 KRT8 KRT6C KRT4 KRT74 LAMB2 MRPL13 DES RPS27 RPL35A KRT10 PCLO ISG15 KRT6A KRT81 TUBB3 FBLN1 OLFM4 RPL34 RPL37 KRT19 RPL39 DCN EPB41L3 CLDN16 CENPA KRT17 RSL24D1 POMZP3 PTPRD LAD1 CLDN1 RPL39L OTOG RPL41 MRPL33 OC90 CLDN23 LAMA4 FRAS1 LUM BGN COL12A1 TINAGL1 MUC3A LAMC2 COL21A1 POSTN HMCN2 NPNT VWA1 THSD4 AGRN ANOS1 EPB41L4B SEPTIN4 APOE LMNA MYBPC1 HLA-DRB1 TPM2 |
| Signaling Receptor Regulator Activity | 63 | KITLG FGF10 CXCL2 CCL22 CLEC11A WNT2 CCL24 CCL7 CCL2 CCL8 CCL1 AREG HBEGF WNT5A INHBA EDN3 EREG PYY CCL25 IL36A MSTN FGF2 CXCL9 FGF7 APP XCL1 LEFTY2 TSLP VEGFC SEMA3D DKK2 LY6E CXCL10 CXCL11 SEMA3E SCG2 CCL13 LYPD6 TAFA5 GDF2 CCL5 IGF1 CTSG TIMP1 IL12B HMGB2 GRN UTS2 RETN EBI3 STC2 IL10 IL21 PROK1 MANF CXCL14 EGFR CLEC12A FLRT2 FST GPNMB SHH CHGB |
| Signaling Receptor Activator Activity | 58 | KITLG FGF10 CXCL2 CCL22 CLEC11A WNT2 CCL24 CCL7 CCL2 CCL8 CCL1 AREG HBEGF WNT5A INHBA EDN3 EREG PYY CCL25 IL36A MSTN FGF2 CXCL9 FGF7 APP XCL1 LEFTY2 TSLP VEGFC SEMA3D CXCL10 CXCL11 SEMA3E SCG2 CCL13 TAFA5 GDF2 CCL5 IGF1 CTSG TIMP1 IL12B HMGB2 GRN UTS2 RETN EBI3 STC2 IL10 IL21 PROK1 MANF CXCL14 EGFR FLRT2 GPNMB SHH CHGB |
| Receptor Ligand Activity | 56 | KITLG FGF10 CXCL2 CCL22 CLEC11A WNT2 CCL24 CCL7 CCL2 CCL8 CCL1 AREG HBEGF WNT5A INHBA EDN3 EREG PYY CCL25 IL36A MSTN FGF2 CXCL9 FGF7 XCL1 LEFTY2 TSLP VEGFC SEMA3D CXCL10 CXCL11 SEMA3E SCG2 CCL13 TAFA5 GDF2 CCL5 IGF1 CTSG TIMP1 IL12B HMGB2 GRN UTS2 RETN EBI3 STC2 IL10 IL21 PROK1 MANF CXCL14 FLRT2 GPNMB SHH CHGB |

**Table:** Enriched Gene Ontology (GO) Analysis Highlighting Upregulated Genes Identified via ShinyGO Database.

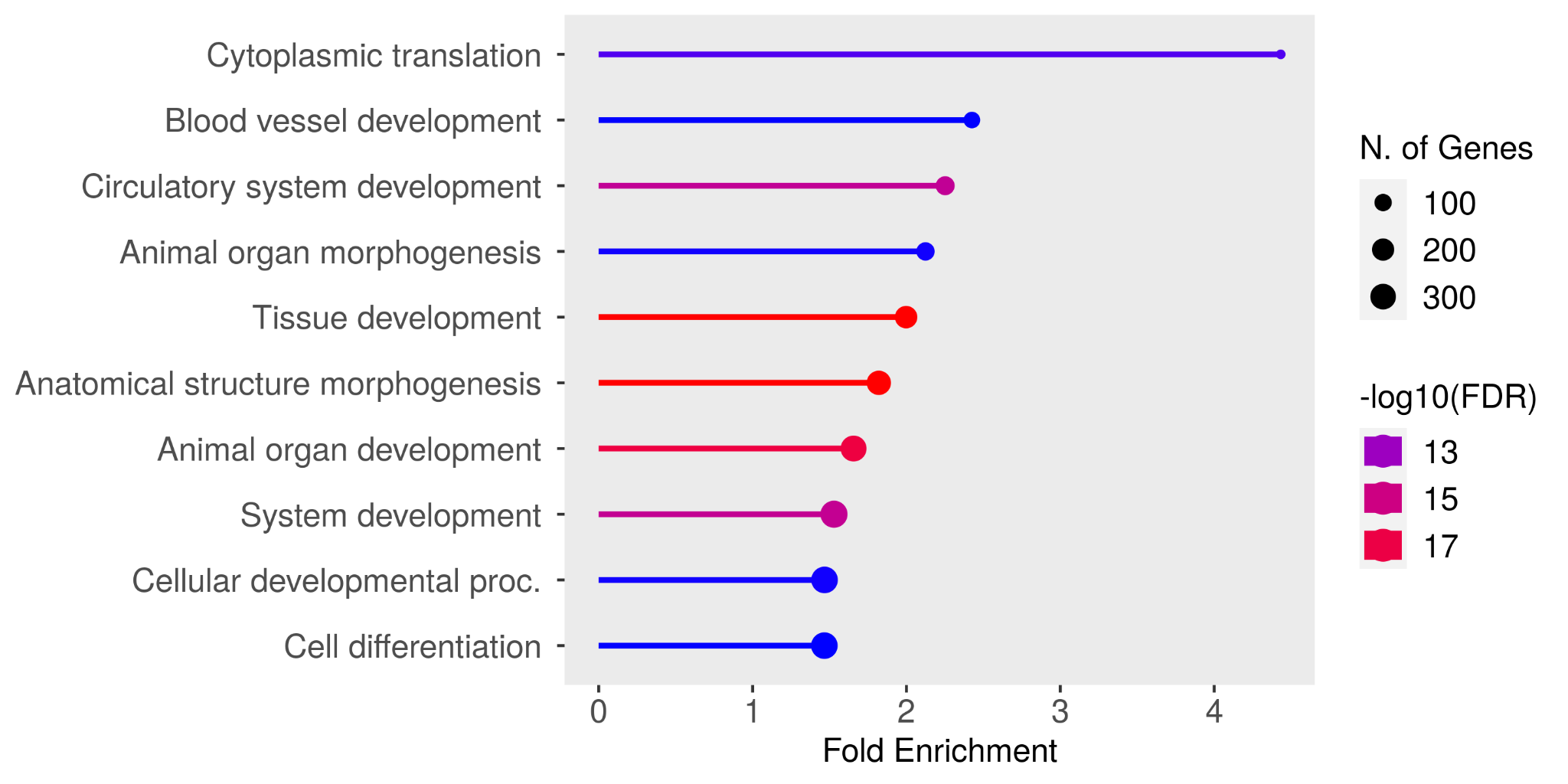

(A)

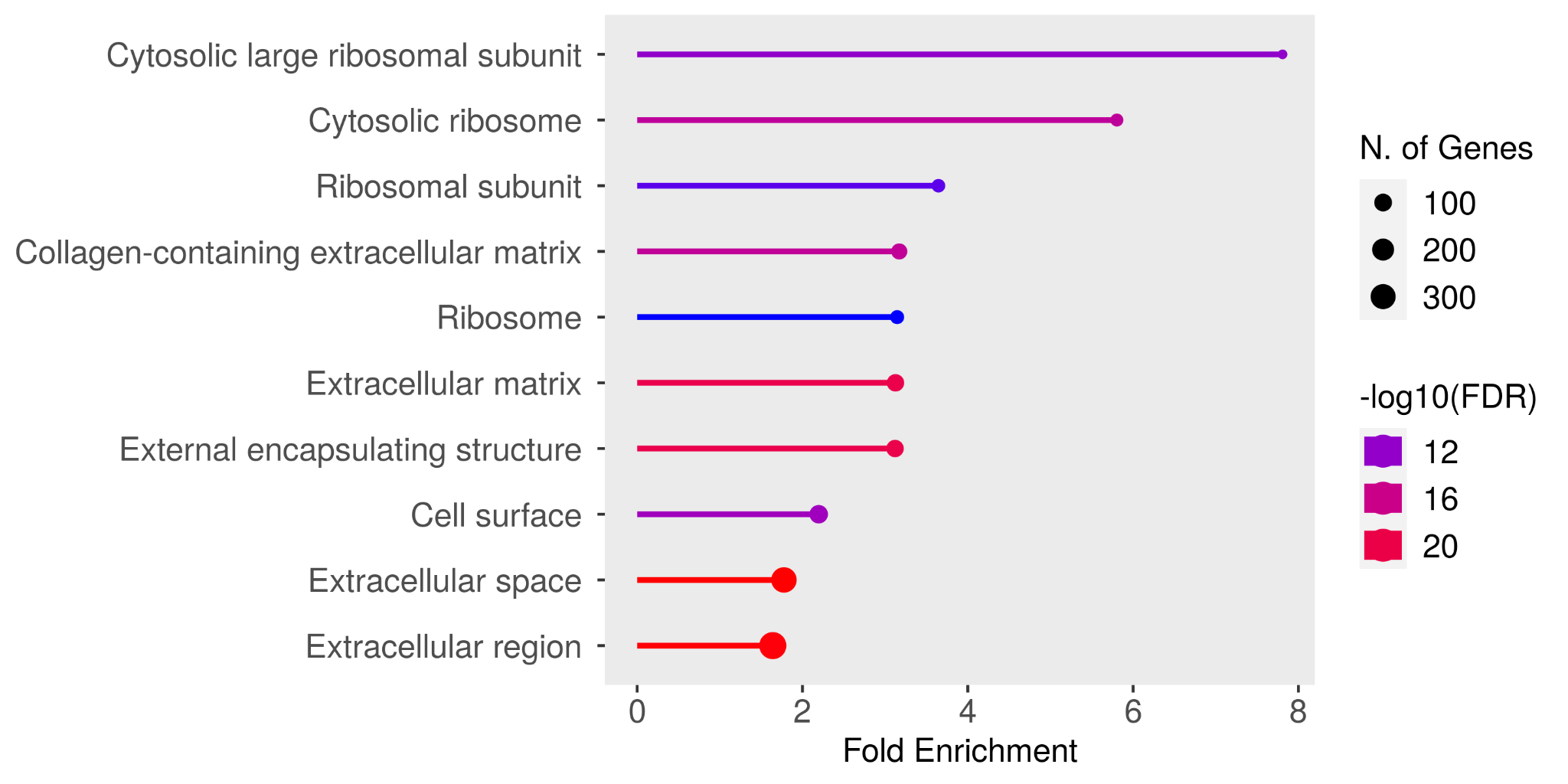

(B)

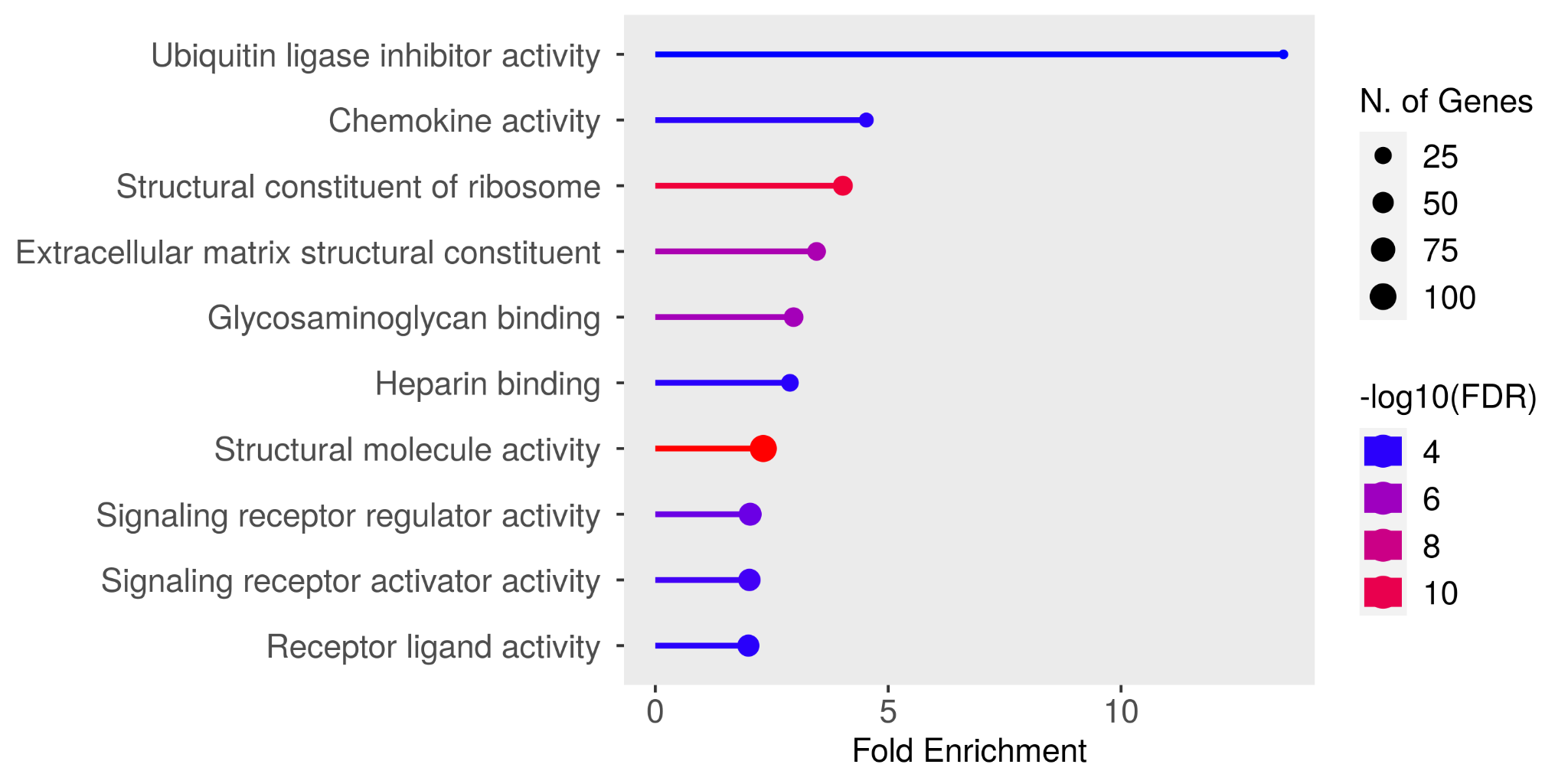
(C)

Fig: Shows the results of a functional enrichment study that focused on upregulated genes, highlighting the enriched (A) biological processes, (B) cellular components and (C) molecular functions associated with upregulated genes. The visualization provides insights into the biological pathways or mechanisms that may be activated or strengthened in response to experimental conditions, aiding in the understanding of gene expression changes in heart failure patients who undergo maintenance hemodialysis
