## Supplementary Table S3 for "Transcriptomic Profiling of Peripheral Blood Mononuclear Cells Reveals Key Molecular Signatures in Chronic Kidney Disease Patients with Heart Failure"

| **Gene Ontology (GO)** | **Number of genes involved (nGenes)** | **Genes** |
| --- | --- | --- |
| **BIOLOGICAL PROCESS** | | |
| protein localization to CENP-A containing chromatin | 14 | H4C8 H4C3 H4C11 H4C14 H4C12 H4C6 H4C13 H4C9 H4C5 H2AC8 H4C4 H2AC4 H4C1 H4C2 |
| Negative reg. of megakaryocyte differentiation | 12 | H4C8 H4C3 H4C11 H4C14 H4C12 H4C6 H4C13 H4C9 H4C5 H4C4 H4C1 H4C2 |
| DNA replication-dependent chromatin assembly | 20 | H4C8 H4C3 H3C12 H4C11 H3C4 H4C14 H4C12 H3C8 H4C6 H4C13 H3C11 H3C1 H4C9 H4C5 H4C4 H3C7 H4C1 H4C2 H3C2 H3C3 |
| DNA replication-dependent chromatin organization | 20 | H4C8 H4C3 H3C12 H4C11 H3C4 H4C14 H4C12 H3C8 H4C6 H4C13 H3C11 H3C1 H4C9 H4C5 H4C4 H3C7 H4C1 H4C2 H3C2 H3C3 |
| nucleosome assembly | 39 | H4C8 H2BC4 H2BC13 H4C3 H4C11 H2BC18 H2BC15 H2BC12L H4C14 H4C12 H2BC14 H2BC6 H4C6 H4C13 H4C9 H2BC3 H4C5 H4C4 H2BC7 H2BC10 H4C1 H4C2 H3C13 H3C12 H3C4 H3C8 H3C11 H3C1 H3C7 H3C2 H3C3 H1-3 H1-1 H1-4 TSPYL6 H1-5 SMYD3 H1-6 H1-2 |
| nucleosome organization | 40 | H4C8 H2BC4 H2BC13 H4C3 H4C11 H2BC18 H2BC15 H2BC12L H4C14 H4C12 H2BC14 H2BC6 H4C6 H4C13 H4C9 H2BC3 H4C5 H4C4 H2BC7 H2BC10 H4C1 H4C2 H3C13 H3C12 H3C4 H3C8 H3C11 H3C1 H3C7 H3C2 H3C3 H1-3 H1-1 H1-4 TSPYL6 H1-5 SMYD3 H1-6 H1-2 SYCP3 |
| chromatin assembly | 39 | H4C8 H2BC4 H2BC13 H4C3 H4C11 H2BC18 H2BC15 H2BC12L H4C14 H4C12 H2BC14 H2BC6 H4C6 H4C13 H4C9 H2BC3 H4C5 H4C4 H2BC7 H2BC10 H4C1 H4C2 H3C13 H3C12 H3C4 H3C8 H3C11 H3C1 H3C7 H3C2 H3C3 H1-3 H1-1 H1-4 TSPYL6 H1-5 SMYD3 H1-6 H1-2 |
| protein-DNA complex assembly | 41 | H4C8 H2BC4 H2BC13 H4C3 H4C11 H2BC18 H2BC15 H2BC12L LINC02218 H4C14 H4C12 H2BC14 H2BC6 H4C6 H4C13 H4C9 H2BC3 H4C5 H4C4 H2BC7 H2BC10 H4C1 H4C2 TAF11L2 H3C13 H3C12 H3C4 H3C8 H3C11 H3C1 H3C7 H3C2 H3C3 H1-3 H1-1 H1-4 TSPYL6 H1-5 SMYD3 H1-6 H1-2 |
| protein-DNA complex subunit organization | 42 | H4C8 H2BC4 H2BC13 H4C3 H4C11 H2BC18 H2BC15 H2BC12L LINC02218 H4C14 H4C12 H2BC14 H2BC6 H4C6 H4C13 H4C9 H2BC3 H4C5 H4C4 H2BC7 H2BC10 H4C1 H4C2 TAF11L2 H3C13 H3C12 H3C4 H3C8 H3C11 H3C1 H3C7 H3C2 H3C3 H1-3 H1-1 H1-4 TSPYL6 H1-5 SMYD3 H1-6 H1-2 SYCP3 |
| chromatin remodeling | 44 | GATAD2B KDM4B H4C8 H2BC4 H2BC13 H4C3 H4C11 H2BC18 H2BC15 H2BC12L KDM4E H4C14 H4C12 H2BC14 H2BC6 H4C6 H4C13 H4C9 H2BC3 H4C5 H4C4 H2BC7 H2BC10 H4C1 H4C2 INO80D H3C13 H3C12 H3C4 H3C8 H3C11 H3C1 H3C7 H3C2 H3C3 H1-3 H1-1 H1-4 TSPYL6 H1-5 SMYD3 H1-6 H1-2 SYCP3 |
| **CELLULAR COMPONENTS** | | |
| CENP-A containing nucleosome | 14 | H4C8 H4C3 H4C11 H4C14 H4C12 H4C6 H4C13 H4C9 H4C5 H2AC8 H4C4 H2AC4 H4C1 H4C2 |
| CENP-A containing chromatin | 14 | H4C8 H4C3 H4C11 H4C14 H4C12 H4C6 H4C13 H4C9 H4C5 H2AC8 H4C4 H2AC4 H4C1 H4C2 |
| chromosome centromeric core domain | 14 | H4C8 H4C3 H4C11 H4C14 H4C12 H4C6 H4C13 H4C9 H4C5 H2AC8 H4C4 H2AC4 H4C1 H4C2 |
| nucleosome | 50 | H4C8 H3C13 H4C3 H3C12 H4C11 H3C4 H3Y2 H4C14 H4C12 H3C8 H4C6 H4C13 H3C11 H3C1 H4C9 H4C5 H4C4 H3C7 H4C1 H4C2 H3C2 H3C3 H1-3 H1-1 H1-4 H2BC4 H2AC20 H2AC21 H1-5 H2BC13 H1-6 H1-2 H2AC13 H2AC11 H2AC7 H2BC18 H2BC15 H2BC12L H2BC14 H2BC6 H2AC12 H2AC15 H2BC3 H2AC16 H2AC8 H2BC7 H2AC4 H2BC10 H2AC17 H2AC18 |
| DNA packaging complex | 50 | H4C8 H3C13 H4C3 H3C12 H4C11 H3C4 H3Y2 H4C14 H4C12 H3C8 H4C6 H4C13 H3C11 H3C1 H4C9 H4C5 H4C4 H3C7 H4C1 H4C2 H3C2 H3C3 H1-3 H1-1 H1-4 H2BC4 H2AC20 H2AC21 H1-5 H2BC13 H1-6 H1-2 H2AC13 H2AC11 H2AC7 H2BC18 H2BC15 H2BC12L H2BC14 H2BC6 H2AC12 H2AC15 H2BC3 H2AC16 H2AC8 H2BC7 H2AC4 H2BC10 H2AC17 H2AC18 |
| protein-DNA complex | 52 | RPA4 GATA1 H4C8 H3C13 H4C3 H3C12 H4C11 H3C4 H3Y2 H4C14 H4C12 H3C8 H4C6 H4C13 H3C11 H3C1 H4C9 H4C5 H4C4 H3C7 H4C1 H4C2 H3C2 H3C3 H1-3 H1-1 H1-4 H2BC4 H2AC20 H2AC21 H1-5 H2BC13 H1-6 H1-2 H2AC13 H2AC11 H2AC7 H2BC18 H2BC15 H2BC12L H2BC14 H2BC6 H2AC12 H2AC15 H2BC3 H2AC16 H2AC8 H2BC7 H2AC4 H2BC10 H2AC17 H2AC18 |
| nuclear chromosome | 28 | SYCP3 SHOC1 IHO1 HUS1B RPA4 INO80D TTN H4C8 H4C3 H3C12 H4C11 H3C4 H4C14 H4C12 H3C8 H4C6 H4C13 H3C11 H3C1 H4C9 H4C5 H4C4 H3C7 H4C1 H4C2 H3C2 H3C3 MSH4 |
| chromatin | 90 | GATAD2B TSPYL6 MXD1 KLF1 INO80D PADI2 H1-3 H1-1 H4C8 H1-4 H3C13 H1-5 H1-2 H4C3 H3C12 H4C11 H3C4 SOX18 H3Y2 H4C14 H4C12 H3C8 H4C6 H4C13 H3C11 H3C1 H4C9 H4C5 H4C4 H3C7 H4C1 H4C2 H3C2 H3C3 SOX10 KDM4B RCOR2 H2BC4 H2AC20 H2AC21 H2BC13 H1-6 H2AC13 H2AC11 H2AC7 H2BC18 H2BC15 H2BC12L H2BC14 H2BC6 H2AC12 H2AC15 H2BC3 H2AC16 H2AC8 H2BC7 H2AC4 H2BC10 H2AC17 H2AC18 NFIX RARB GATA1 OVOL3 HOXA2 SOX6 BACH2 OTX1 FOXO3 MXI1 BARHL1 NPAS1 RHOXF2 GCM1 ESR2 NKX6-2 FOXO1 NR3C2 EBF1 NKX3-1 ETV4 ZBED2 FOXE1 DMRTC1B OLIG2 POU5F1B MEF2B POU5F2 HSFX3 SCRT2 |
| Intrinsic component of plasma membrane | 116 | DRD4 TRPM5 ASIC4 TRPC5 HRH3 HAS3 HAS1 RNF43 PRPH2 ELAPOR1 TRPM6 CHRNA2 HCRTR1 KCNS1 LRRN4 AVPR2 ADORA2A LRRC4 OPN1SW TSPAN16 SLC6A8 ADGRE3 CHRND HTR2B PCDH8 ITGA11 DUOX1 SLC28A2 SLC38A4 SLC39A5 IGF1R KCNH1 SLC26A1 KIRREL3 EPHB1 HTR6 CHRNB2 GRIN2C BSND FCGR3B F2RL2 SLC16A4 TSPAN5 GYPA ALK GPR22 RXFP4 KCNK7 ADGRG2 KCNG2 ADGRG3 KCNH7 PMEL ZACN RHD SHISA7 NLGN3 SLC6A9 AJAP1 CHRNG GYPE GPR52 IL10RB GYPB PCDHGB1 LYPD4 SLC4A1 CDHR2 ATP8B1 RAMP3 KCNJ2 OSMR ADIPOR1 CA4 HJV TMEM266 LRRC8E ENPP7 EPGN CNTN2 P2RX2 IFITM5 SLC12A5 MMP24 ACKR2 KCNJ9 TMC4 RAB26 SLC19A1 SLC22A1 TMPRSS9 NRXN1 UNC5A CACNA1C TNFSF4 MME AGER TNFRSF9 SPTB TACR2 SLC10A1 CRHR2 TACR1 MLANA FFAR2 PODXL LYVE1 GYPC SORL1 KCNJ15 SLC13A4 SLC35G2 CXCR2 IL1RAP SLC22A4 HCAR3 |
| extracellular region | 242 | SLC4A1 TMEM132A BAIAP2L1 CP PRSS8 GSTO2 CDHR2 SLC12A1 MCAM LAMB1 IGFALS SERPIND1 SEC14L2 UPB1 ACTR3C FAM20A VTN THBS4 HYAL1 ANGPTL1 ELAPOR1 OLFML3 PADI2 MUC5B RARRES1 PPL FABP3 SDCBP2 TGM3 LRRN4 MMP24 PZP STEAP4 PODXL SHBG PLVAP GDF15 MYH10 CHI3L1 MYH11 LYVE1 ANGPTL2 TUBB2A SORL1 LOXL4 SLC39A5 SLC5A2 SELENBP1 CRB2 SLC5A10 ABI3BP EPHB1 TTN H4C8 PIP NAPEPLD PDZK1IP1 ALPL FCGR3B CTHRC1 MFAP4 LDHAL6A STX3 TBC1D21 CA4 UBXN6 KRT1 ANGPTL4 RPS9 ALK ADGRG2 A2M KCNG2 H2BC4 H3C13 H2AC20 H2AC21 H2BC13 ZNF177 MME H2AC13 H2AC11 H2AC7 H4C3 H3C12 H4C11 H3C4 H2BC18 MBD5 PRRC2A PRSS1 GPX3 ORM2 H2BC15 UBE2V1 TRIM75 MGAM HPR H4C14 H4C12 H2BC14 H3C8 H2BC6 H4C6 H2AC12 H4C13 H2AC15 H3C11 H3C1 H4C9 H2AC16 H4C5 H2AC8 H4C4 H2BC7 PKD1L3 H3C7 H2AC4 H2BC10 H4C1 H2AC17 H4C2 H3C2 H3C3 H2AC18 CRLF1 ANGPT2 PGC OXT PRSS33 DKK4 CGB3 TNFSF4 SFRP5 COL10A1 PI3 CXCL6 GNRH2 LRRC17 LOXL1 CBLIF SFRP2 SCUBE3 IFNK GHRL CLEC18A CLEC18C AGRP FNDC5 WNT4 INHBB TGFA CXCL1 EVA1C SFTPB ANGPTL7 IFNB1 TAC4 EPGN OLFML1 LYG2 SERPINA11 CXCL17 CR1L HLA-F CCL27 LINGO3 DEFB131B CNTF SERPINE3 LYPD8 CCN5 NTN1 RS1 HAS3 PYCR3 APC2 CHRNA2 SNCA EPB41 HJV ALKAL2 CKLF ERVH48-1 H2BC12L NOTCH2NLB MMP25 METTL24 FGF22 CMTM1 SMPD3 CEMIP SCG3 CNTNAP3 RLN1 ENDOU IL1R2 INSL6 PRB2 SPINK4 ADGRE3 PDZD2 CNMD CCN3 ADAMTS17 ADCY10 ROPN1L SPINK7 KIRREL3 VWA5B1 NTN3 IL24 C1QTNF7 PROK2 SCRG1 KIF27 TUB VWCE NRTN PHOSPHO1 NXPH3 DEFB108B PMEL EMID1 OVCH1 TREML4 QRFP PLAC9 IL1RAP AGER COL6A6 C1QTNF9 TNFRSF6B PRSS51 NOTCH2NLA EDDM13 C1QTNF3-AMACR F2RL2 PLET1 ATG7 SPRN LYPD4 |
| **MOLECULAR FUNCTIONS** | | |
| interleukin-8 binding | 3 | CXCR1 CXCR2 A2M |
| hyaluronan synthase activity | 3 | HAS3 HAS1 HYAL1 |
| structural constituent of chromatin | 50 | H1-3 H1-1 H4C8 H1-4 H2BC4 H3C13 H2AC20 H2AC21 H1-5 H2BC13 H1-6 H1-2 H2AC13 H2AC11 H2AC7 H4C3 H3C12 H4C11 H3C4 H2BC18 H2BC15 H2BC12L H3Y2 H4C14 H4C12 H2BC14 H3C8 H2BC6 H4C6 H2AC12 H4C13 H2AC15 H3C11 H3C1 H4C9 H2BC3 H2AC16 H4C5 H2AC8 H4C4 H2BC7 H3C7 H2AC4 H2BC10 H4C1 H2AC17 H4C2 H3C2 H3C3 H2AC18 |
| protein heterodimerization activity | 52 | BCL2L1 KRT1 NPAS1 H4C8 SLC51A H2BC4 H3C13 H2AC20 H2AC21 H2BC13 H2AC13 H2AC11 H2AC7 H4C3 H3C12 H4C11 H3C4 H2BC18 H2BC15 H2BC12L LINC02218 H3Y2 H4C14 H4C12 H2BC14 H3C8 H2BC6 H4C6 H2AC12 H4C13 H2AC15 H3C11 H3C1 H4C9 H2BC3 H2AC16 H4C5 H2AC8 H4C4 H2BC7 H3C7 H2AC4 H2BC10 H4C1 H2AC17 H4C2 TAF11L2 H3C2 H3C3 H2AC18 SDCBP2 AOC3 |
| structural molecule activity | 83 | COL10A1 RPS9 TUBA3D LAMB1 CRYBA1 PPL TUBB2A KRT1 CSRP2 MBP TUBA4B PI3 TGM3 TTN SPTB H1-3 H1-1 H4C8 EPB41 H1-4 CLDN20 SNTG2 H2BC4 H3C13 H2AC20 H2AC21 H1-5 H2BC13 H1-6 H1-2 H2AC13 H2AC11 H2AC7 H4C3 H3C12 H4C11 H3C4 H2BC18 CLDN9 H2BC15 H2BC12L H3Y2 H4C14 H4C12 H2BC14 H3C8 H2BC6 H4C6 H2AC12 H4C13 H2AC15 H3C11 H3C1 H4C9 H2BC3 H2AC16 H4C5 H2AC8 H4C4 H2BC7 H3C7 H2AC4 H2BC10 H4C1 H2AC17 H4C2 H3C2 H3C3 H2AC18 MYH11 PANX2 VTN ANK1 CHI3L1 ABI3BP CTHRC1 MFAP4 COL6A6 MYOT MYBPH EPB42 SEPTIN5 ACTL7A |
| protein dimerization activity | 75 | BCL2L1 TBC1D22A FECH UPB1 BNIP3L PADI2 SDCBP2 GDF15 PDLIM4 CBS KRT1 TMEM266 MME ATG7 SMIM1 MXD1 DGAT2 PRPH2 MXI1 NPAS1 THRSP H4C8 SLC51A H2BC4 H3C13 H2AC20 H2AC21 H2BC13 H2AC13 H2AC11 H2AC7 H4C3 H3C12 H4C11 H3C4 PLN H2BC18 OLIG2 MEF2B H2BC15 H2BC12L LINC02218 H3Y2 H4C14 H4C12 H2BC14 H3C8 H2BC6 H4C6 H2AC12 H4C13 H2AC15 H3C11 H3C1 H4C9 H2BC3 H2AC16 H4C5 H2AC8 H4C4 H2BC7 H3C7 H2AC4 H2BC10 H4C1 H2AC17 H4C2 TAF11L2 H3C2 H3C3 H2AC18 SOX6 SLC4A1 AOC3 INHBB |

**Table:** Enriched Gene Ontology (GO) Analysis Highlighting Upregulated Genes Identified via ShinyGO Database.

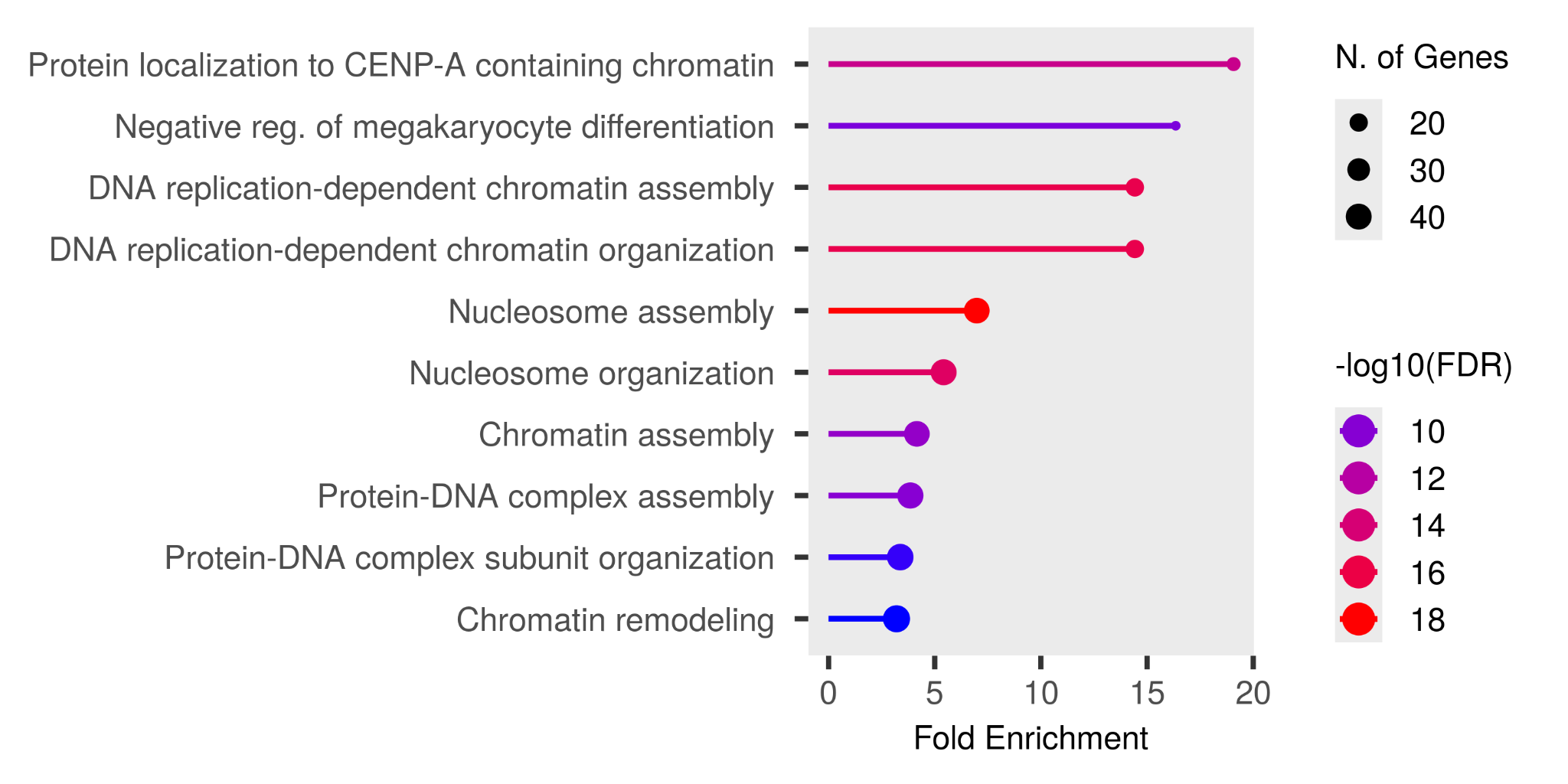

(A)

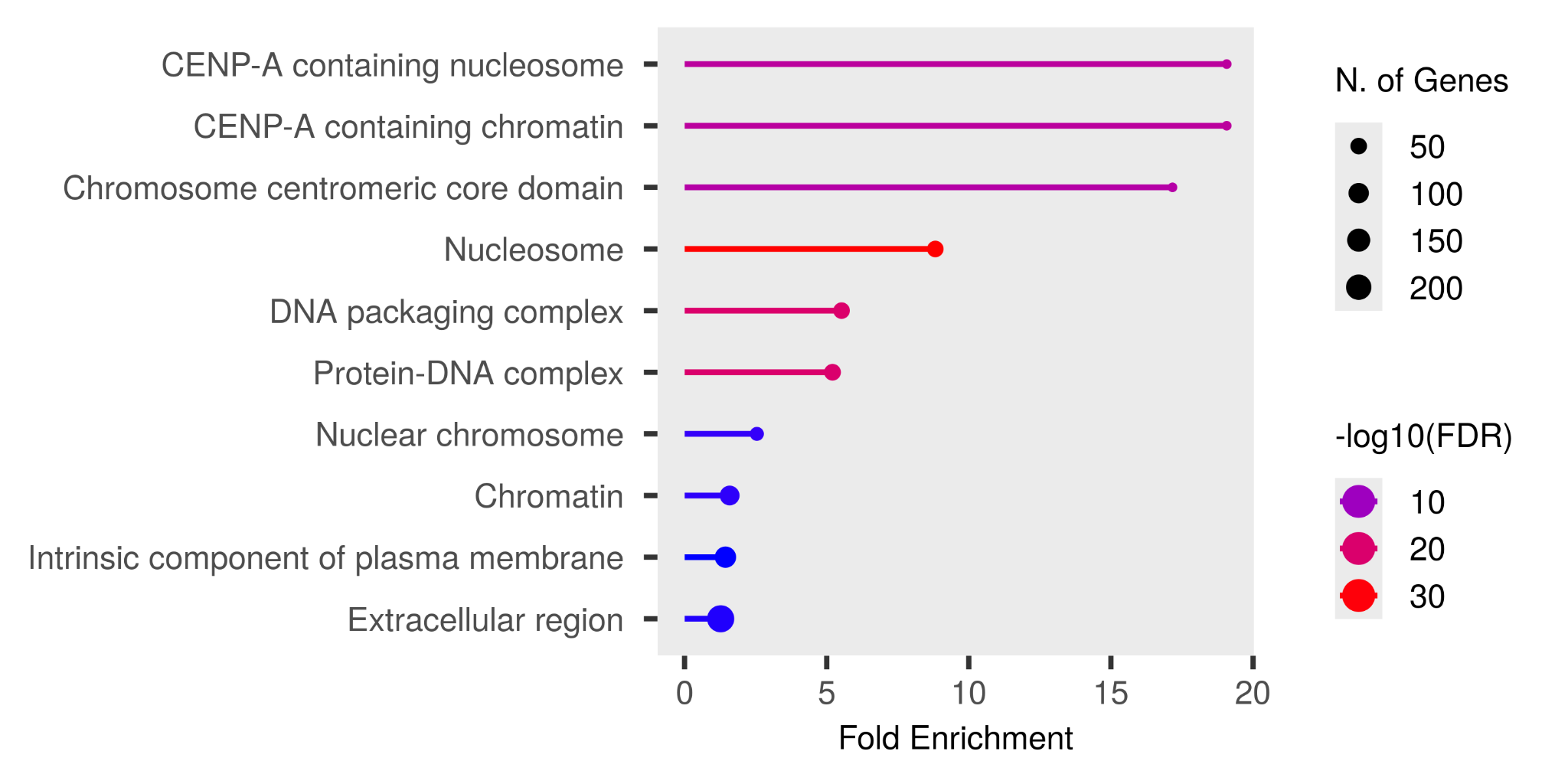

(B)

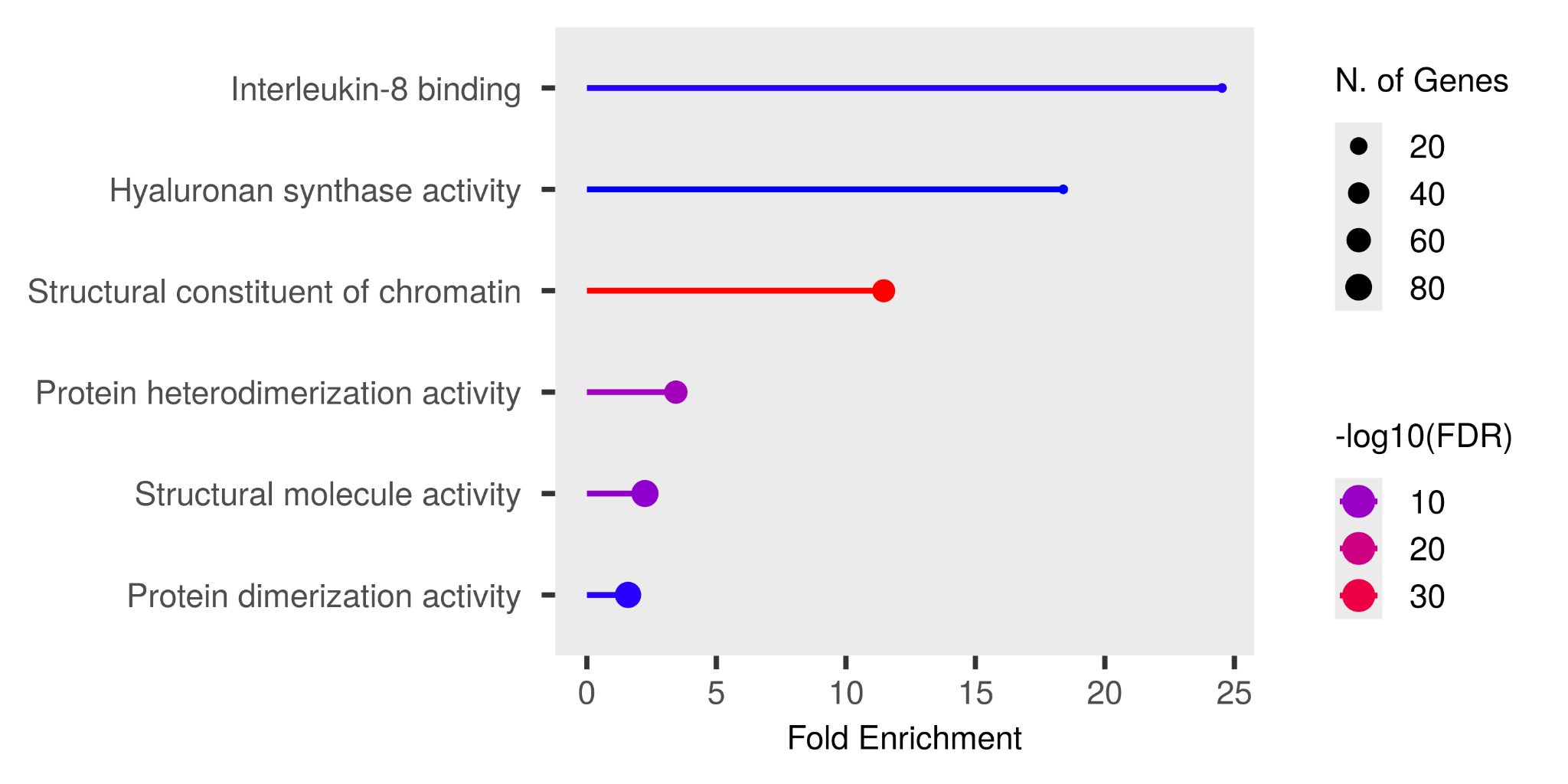

(C)

Fig: Shows the results of a functional enrichment study that focused on downregulated genes, highlighting the enriched (A) biological processes, (B) cellular components, and (C) molecular functions associated with upregulated genes. The visualization provides insights into the biological pathways or mechanisms that may be activated or strengthened in response to experimental conditions, aiding in the understanding of gene expression changes in heart failure patients who undergo maintenance hemodialysis.
